# Immunity acquired by a minority active fraction of the population could explain COVID-19 spread in Greater Buenos Aires (June-November 2020)

**DOI:** 10.1101/2021.12.21.21267955

**Authors:** Gabriel Fabricius, Rodolfo A. Borzi, José Caminos, Tomás S. Grigera

## Abstract

The COVID-19 pandemic had an uneven development in different countries. In Argentina, the pandemic began in march 2020 and, during the first 3 months, the vast majority of cases were concentrated in a densely populated region that includes the city of Buenos Aires (country capital) and the Greater Buenos Aires area that surrounds it. This work focuses on the spread of COVID-19 between June and November 2020 in Greater Buenos Aires. Within this period of time there was no vaccine, basically only the early wild strain of SARS-CoV-2 was present, and the official restriction and distancing measures in this region remained more or less constant. Under these particular conditions, the incidences show a sharp rise from June 2020 and begin to decrease towards the end of August until the end of November 2020. In this work we study, through mathematical modelling and available epidemiological information, the spread of COVID-19 in this region and period of time. We show that a coherent explanation of the evolution of incidences can be obtained assuming that only a minority fraction of the population got involved in the spread process, so that the incidences decreased as this group of people was becoming immune. The observed evolution of the incidences could then be a consequence at the population level of lasting immunity conferred by SARS-CoV-2.

## I. INTRODUCTION

Since the COVID-19 pandemic began in late 2019 in Wuhan (China), the disease has become a serious public health problem globally. Understanding the transmission dynamics of COVID-19 is a matter as complex as it is important, since any advances facilitate the taking of adequate control measures.^1^ Although the transmission of any infectious disease is complex, in the case of COVID-19, several specific features of the disease make it difficult to characterise and control the contagion process. The possibility of contagion before the appearance of symptoms, the existence of asymptomatic individuals, the diversity in the symptoms and the heterogeneity in the immune response observed in different individuals, are just some of the characteristic elements that have become known as the pandemic moved along.

One of the most controversial aspects of the disease is the issue of the immunity generated by SARS-CoV-2 in individuals. The mechanisms of natural immunity and the degree of protection against new infection they eventually confer are subject of intense research.^2^ At this point, it is quite clear that natural immunity protects against disease, but the degree of protection obtained against contagion is not so evident.^2–5^ The epidemiological impact of individual immunity is even less clear. This is difficult to assess because every epidemiological observable (such as reported cases or serological studies, for example) is affected by a multiplicity of factors, such as effectiveness of health system surveillance or uncertainties in antibody test results. In some cases, the study of the evolution of the pandemic in certain regions at definite times, offers the possibility of exploring the relationship of some of these factors with the temporal evolution of the reported cases. This is what we set to do here, taking advantage of the particular conditions that occurred in a region of Argentina during a six-month period within the pre-vaccination era.

The evolution of the pandemic has produced rising and falling incidence curves in every region, but the reasons behind this behaviour are often very different. For example, the first outbreak observed in Italy between March and June 2020 can be explained by assuming that at the beginning of March the disease spread freely with a high *R*_0_. Severe restrictions were imposed, causing the value of *R*_0_ to drop violently below 1, and a consequent decrease in daily cases to values insignificant compared to those at the peak.^6^ The evolution of cases in Greater Buenos Aires (GBA), Argentina, between June and the end of November 2020 (Fig. 1) is qualitatively similar, but requires a very different explanation. In Argentina, severe isolation measures were established a few days after the appearance of the first cases, when there was practically no community transmission; these measures were then gradually relaxed over time, and never hardened again. At the beginning of June there was a sustained community transmission in all GBA districts, with an average incidence of between 4 and 5 cases per day per 100,000 inhabitants, but no new measures were taken. Thus, the sustained fall of incidence observed since the end of August in Argentina must have a different origin than in the Italian case. Although it is possible that some health measures, such as the use of the face mask or the protocols in hospitals, had been improving over time, it is difficult to attribute the drop to these reasons. Indeed, public data^7,8^ (Fig. S1) show increasing mobility in all spaces, even as the incidence continued to decrease. A plausible hypothesis is that this sustained decrease was due to the acquisition of immunity. In this work we explore, through mathematical modelling, under what conditions the fall in incidences that occurs between the end of August and November (which is a phenomenon observed in the 24 GBA districts, Fig.S2) can be explained from the acquisition of immunity by vast sectors of the population. The results of our simulations show that there are several scenarios compatible with the available epidemiological information on the spread of COVID-19 in GBA which account for the observed dynamic evolution of the incidences, as long as it is assumed that the transmission process was basically caused by a minority but active fraction of the population that was acquiring immunity.

**FIG. 1.**
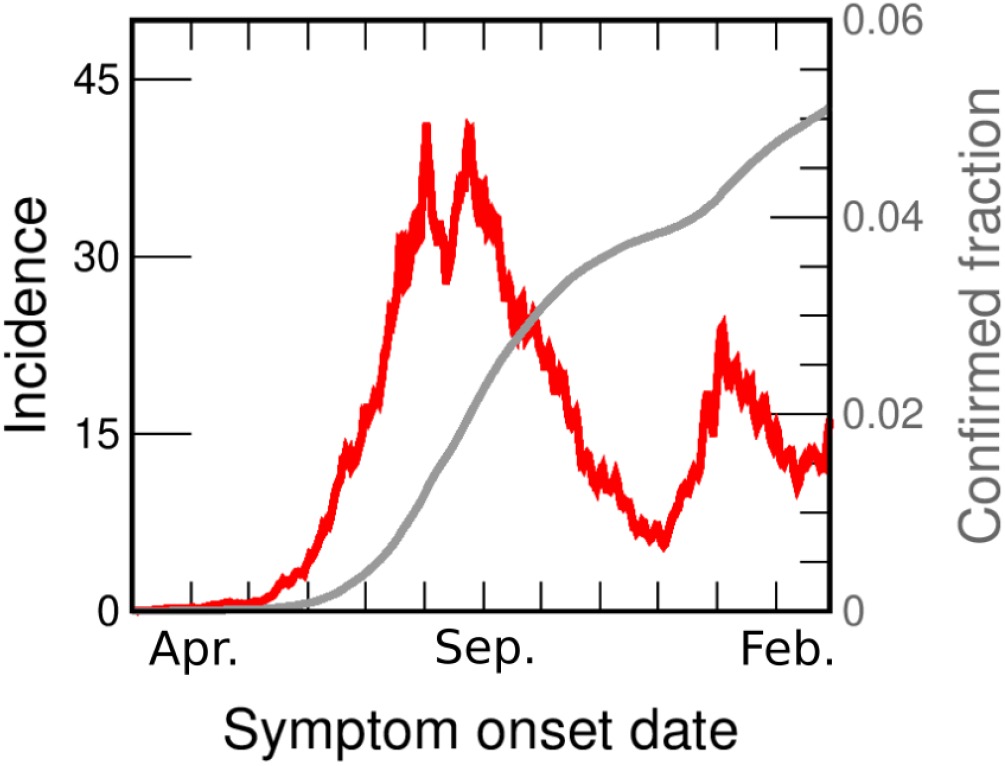
COVID-19 incidences in the Greater Buenos Aires (GBA) from March 2020 to February 2021. Data are consigned in cases per day per 100, 000 inhabitants, (red line, left axis). We also show the fraction of the population reported as confirmed case (gray line, right y-axis). Both curves are drawn as a function of the symptoms onset date (SOD), estimated from reported data.^9^ Details of the incidences computation are given in the Supplementary Material (SM), section I.B.

## II. MATERIALS AND METHODS

### A. Demographical, epidemiological and mobility data

Located in Buenos Aires province, the Greater Buenos Aires (GBA) is made up of the 24 districts closest to Buenos Aires city (CABA) with a total population of 9,916,715 inhabitants according to the last census carried out in 2010. Except for La Matanza, which has a population of 1,775,816 inhabitants, all the other districts have less than 650,000 pop. There are 4 districts (San Fernando, Ezeiza, Ituzaingó and Hurlingham) with less than 200,000 pop., 10 districts (José C. Paz, Vicente López, San Miguel, San Isidro, Esteban Echeverría, Morón, Malvinas Argentinas, Berazategui, Tres de Febrero and Avellaneda) that have between 250,000 and 350,000 pop., 3 districts (Tigre, General San Martín and Florencio Varela) between 350,000 and 450,000 pop., 3 districts (Moreno, Lanús and Merlo) between 450,000 and 550,000 pop. and 3 districts (Almirante Brown, Quilmes and Lomas de Zamora) between 550,000 and 650,000 pop. The projected GBA population for July 2020 is 11,264,104,^10^ therefore, in order to calculate the incidences, we will correct all the populations by a multiplicative factor of: 1.136 = 11,264,104 / 9,916,715.

The first confirmed case of COVID-19 in Argentina was on March 3rd, 2020. On March 16th, schools and non-essential activities were called off in large cities. On March 20th, a lock-down (Preventive and mandatory social isolation or ASPO in Spanish) was decreed throughout the country, and proceeded from that moment on, with changes to different phases in different regions. On June 4th, 18 provinces discontinued their lock-down, but the GBA was among the regions that did not. Strictly speaking, the GBA was in lock-down from March 20th to November 9th, when it was replaced with social distancing measures (preventive and mandatory social distancing or DiSPO in Spanish). That is, throughout the period to be studied here a lock-down was officially in place.^11^ However, communication and actual enforcement of the measures was variable through the period, and so was compliance. Mobility data from Google^7^ (Figure S1(a)) indicates that mobility in supermarkets, jobs and transport stations began to increase sharply just after the ASPO was decreed until the beginning of June. From June, these indices remained constant or increased at a much slower rate.

Fig. 1 shows the incidence in GBA vs. date of onset of symptoms (the same data for individual districts is in Fig. S2). From the beginning of June, the incidence in GBA shows a sustained growth (at approximately constant rate) for 2 months, from between 4 and 5 daily cases per 100,000 pop. to 40 daily cases / 100,000 pop. Then for the next 20 days it oscillates around 35 daily cases / 100,000 pop. and finally begins to fall steadily until the end of November. The rate of decrease is also approximately constant but lower than the initial increase rate. Although the evolution of incidences shows some peculiarity in each individual district, the mentioned qualitative features are the same across the whole GBA region. In most districts the rise and fall times coincide with the global GBA curve, and in some districts (such as Almirante Brown, Berazategui or Morón) even the absolute values are very similar to the GBA case throughout the period.

The incidences estimated from officially reported cases are a lower bound for the actual incidences of COVID-19. The causes of under-reporting are varied and most likely originate in cases that were asymptomatic or with weak symptoms, although there may be other reasons, such as those concerning the efficiency of the detection system. It is very difficult to estimate the number *k* of actual cases per reported case. The literature estimates vary widely, according to the criterion adopted and the stage of the pandemic.^12–17^ We discuss under-reporting for GBA in the Supplementary Material (SM: section I.D) and conclude that a plausible range for the *average* value of *k* in the studied period is between 3 and 6. However, it is important to keep in mind (as we discuss later) that under-reporting could vary somewhat over time, and also be different in different districts.

### B. Mathematical modelling

In a subject with the complexity of the spread of COVID-19, it is impossible to consider a mathematical model that takes into account all the elements and the heterogeneity present in the problem. The spirit of our modelling is to take into account the central ingredients, assume certain hypotheses (based on a set of evidences) and then use the model as an exploration tool. In particular, we are interested in exploring under what conditions the model reproduces the increase in incidences by a factor of approximately 8 after approximately 70 days and the subsequent decrease by a similar factor in a slightly longer time, both observed in the dynamic evolution of the reported incidences (Fig.1 and Fig.S2). Given the uncertainties in the knowledge of the transmission process and the ignorance of the cases of COVID-19 that really existed in the analysed period (of which the reported cases are only a fraction), we do not seek to obtain the set of model parameters that best fits the reported cases, but to evaluate the degree of plausibility of different scenarios.

#### 1. Assumed hypotheses

To build our model we assume the following:

1. All infected individuals go through a latency period after which they become contagious. We not distinguish between the contagion capacity of symptomatic and asymptomatic individuals, nor will we take into account whether some were isolated and others not: all infected individuals infect equally but the time each individual is contagious can vary and is described by a probability distribution.
2. When infected individuals recover, they acquire immunity lasting at least 6 months (the period we attempt to model^18^).
3. The structure of social contacts through which contagions occur stayed the same in GBA between June and November 2020. This is probably not strictly true, but nevertheless a reasonable working hypothesis.
4. Only a fraction *f*_*C*_ of the individuals participated in the contagion process. This hypothesis is based on the situation that occurred in Argentina, where some individuals were very little exposed to contagion (because they performed virtual tasks, and when they went out they took care of themselves using face mask and respecting the decreed social distancing) while others suffered a much higher degree of exposure (because they did not comply with the standards of care in force, or due to their work situation). There were probably several intermediate behaviours but we will assume as a working hypothesis that the population is divided into two types of individuals: those who participate in the contagion process and those who do not.
5. In the period studied (June-November 2020), the strongest contacts occurred at home, the next strongest among groups of close families, then among the neighbourhood and so on, decreasing as the spatial scale increases. This is justified given that there were still serious restrictions on allowed activities.
6. The epidemic “ran by itself” within each district of GBA from June on, when community transmission was already occurring in most neighbourhoods. That is, we neglect effects due to cases imported from other districts or from abroad. This would not be justified in the previous months, in which the influence of imported cases from abroad or from the city of Buenos Aires may have played an important role. The simulations we have carried out to validate this hypothesis are shown in the SM (section III.B.2).

#### 2. Model Description

We consider a stochastic epidemiological model where a susceptible individual, once infected, goes through the chain of states *S* → *E*_1_ → *E*_2_ → *I*_1_ → *I*_2_ → *R*, corresponding respectively to *susceptible*, two stages of *exposed*, and two stages of *infected* individuals. (see Fig. 2). The population is made up of *N* individuals where the individual *j* is in the epidemiological state *X*_*j*_. The state of the system at a given instant is defined by X = (*X*_1_, *X*_2_, …*X*_*N*_). **X**(*t*) is a stochastic variable that evolves in time according to the Markov process defined by the transitions indicated in Fig. 2.

**FIG. 2.**
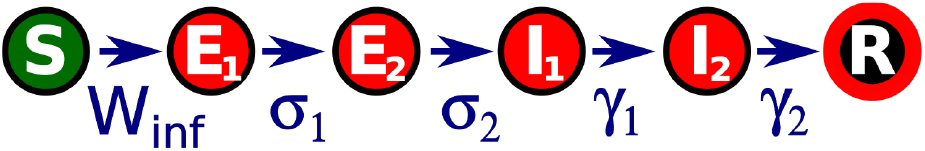
Epidemiological model used in this work. A given individual is described by its epidemiological state *X*. A susceptible individual (*X* = *S*) may became infected in contact with an infectious individual (*I*_1_ or *I*_2_, which represent two stages at which the individual can infect) with a probability rate *W*_inf_. *E*_1_ and *E*_2_ are latency states (the *exposed* individual is infected but cannot yet infect). The transitions from *E*_1_ to the successive states occur randomly with constant probability rates *σ*_1,2_ and *γ*_1,2_. The final state (*X* = *R*) represents a recovered, immune individual. We assume that there is no conversion from *R* to *S* during the period of our study (possible effects of this not being true are discussed in the SM (section III.B.1). Two epidemiological states have been chosen to describe the latency phase and another two for the infectious phase so that the model reproduces distributions of latency and infection times compatible with those reported in literature and available epidemiological data, see SM (section II.B). Using more than one state for the same epidemiological phase is known to produce more realistic transition dynamics,^19^ in particular two classes have also been used to model COVID-19 by other authors.^20^ *W*_inf_ is not constant, but depends on the instantaneous configuration of infected individuals (see text).

The probability of contagion per unit of time of individual *j*, 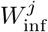 is determined by the assumed social structure. We use a hierarchical contact structure like the one schematised in Fig. 3. This allows considering a certain spatial heterogeneity in the spread of the disease compatible with the mobility restrictions in force in the period studied. Each individual is associated with a group of a given level. Within each level *l* (household, building, neighbourhood and so on) an individual interacts homogeneously with the other individuals in the same group with the same effective rate of contacts per unit of time. We indicate with *l* = 1, 2, …, *L* the level in the group hierarchy. An individual *j* belongs to group *ν*_*l,j*_ at level *l*, which has 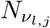 members. The probability of transition per unit of time for the infectious process is given by

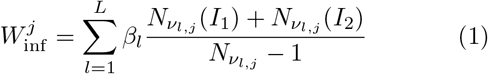

where 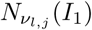 and 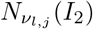 are the numbers of individuals of group *ν*_*l,j*_ in states *I*_1_ and *I*_2_ respectively. The parameters *β*_*l*_ are associated with the probability of contacts per unit of time between individuals within the level *l*. In addition, according to hypothesis 4 above, some families (groups with *l* = 1, level “household”) are randomly marked “inactive” (i.e. they have no social contact) and do not participate in the contagion dynamics even if they are in state *S*. These families can eventually become active at a later stage of the dynamics.

**FIG. 3.**
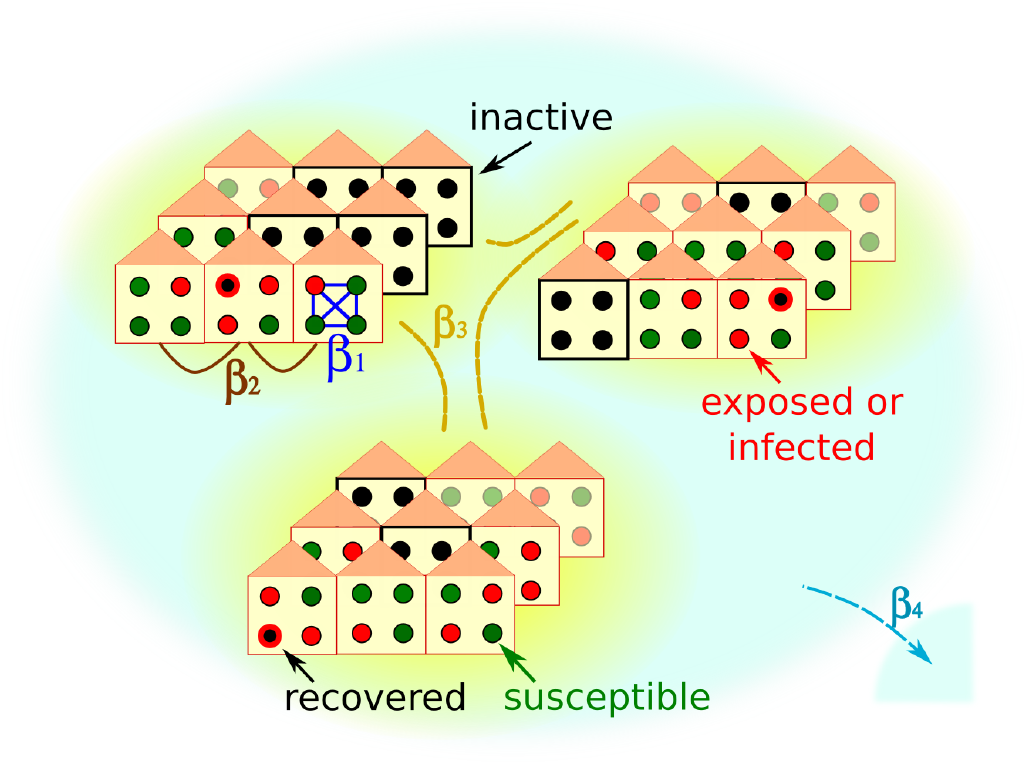
Schematic representation of our hierarchical model. Individuals (coloured circles representing states as in Fig. 2) are hierarchically grouped in families which belong to buildings, neighbourhoods, etc.. Black circles are either recovered (red edge) or inactive. In either case they are inert as regards their capacity to infect or become infected. Recovered individuals remain in this state for the whole simulated period, while some of the inactive may be activated (turning them to susceptible) in order to simulate a behavioural change in those individuals. The fraction, *f*_*C*_, of inactive individuals is a parameter of the model and fixed at the beginning. Inactive individuals always belong to isolated families (marked by black borders and fully occupied by inactive individuals), chosen randomly within the different neighbourhoods and towns. Some spatial structure is taken into account by the hierarchical nature of the model. The contact rate for individuals of the same household (*β*_1_, marked in blue) is assumed stronger than for individuals from different households (*β*_2_, brown). Individuals from different *neighbourhoods* interact with an even smaller rate (*β*_3_, yellow), while people from different *towns* (the next hierarchical level) interact with *β*_4_ (light blue).

We will take 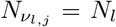, depending only on the level (i.e. all families, neighbourhoods etc. are all the same size). Note however that since the choice of which families will be inactive is random, the number of active individuals will be different for different groups at the same level for *l >* 1, and therefore the effective contact rates will fluctuate around a mean value 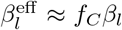 for *N*_*l*_ ≫ 1, where *f*_*C*_ is the fraction of active individuals, see SM (section II.A). To simplify matters, we will generally refer to a given level using the number of individuals in a given group of this level rather than the index *l*. For instance, a level with groups of *N*_*l*_ = 20000 individuals will be indicated as a “n2E4 neighbourhood”. A group of the first level is usually mentioned as a family or household. Stochastic simulations are performed with the Gillespie algorithm.^21^ Simulation software is available for download at the GitHub repository (see Data Availability section). The incidence Inc(*t*) (in cases per day) is computed as the number of individuals whose state changes from exposed to infected (*E*_2_ → *I*_1_) during day *t*, and the reproductive ratio, *R*(*t*), at day *t* as

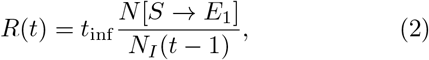

where *t*_inf_ is the mean duration of infection, *N* [*S* → *E*_1_] is the number of individuals whose state changes from susceptible to exposed between day *t* − 1 and day *t*, and *N*_*I*_ (*t* − 1) is the total number of infected individuals (in states *I*_1_ or *I*_2_) at day *t* − 1.

## III. RESULTS

For the results presented in this section, we take *σ*_1_ = *σ*_2_ = 0.66667 1/day which corresponds to an average latency time of 3 days, *γ*_1_ = *γ*_2_ = 2*γ* = 0.28571 which corresponds to a mean time of infection *t*_inf_ =1*/γ*=7 days, and *β*_1_=0.214 1/day for the rate of household contacts. In the SM (section II.B) we provide a justification for the choice of these values based on several studies and GBA-epidemiological data.^9,22–28^ For an analysis of the robustness of the results under changes in the parameters within a plausible range of values, see SM (section III.A).

To motivate the need for the spatial model we have introduced, we begin by studying the evolution of the incidences using the much simpler uniform mixing approach. Figure 4 shows the incidence as a function of time under uniform mixing for two values of *R*_0_. What we observe is that for *R*_0_=1.45, the incidence grows by a factor of 8 until reaching the maximum in a time of 70 days (as observed in the incidences obtained from reported cases of the GBA, Figure 1). However, the absolute values obtained are almost 20 times greater than those reported, resulting in 55.0% of the population infected at the end of the epidemic (a much higher proportion than the 11.6% obtained from serological studies^30^). On the other hand, for *R*_0_=1.17 the incidence obtained from the model takes values around 3 times those reported (which are within the plausible range), but the development time of the epidemic turns out to be greater than one year (more than twice the observed time). It is clear that the homogeneous mixing approach gives a description of the development of the epidemic that does not even approximate the orders of magnitude involved, pointing that a drastic change in this approach is necessary. One possibility (and one of the key points in this work) is to assume that only a small fraction of the population participates in the contagion process; this could explain how a maximum in the incidence curve could be attained in a relatively short time without a huge fraction of the population getting the disease. Instead, the peak would occur when a relevant fraction of the most *active* (regarding their chances of getting and spreading the illness) population is infected. For example, assuming that 25% of the population is active and contacting each other homogeneously while the rest remain inactive, for a value of *R*_0_=1.45, the number of cases would be reduced to 25% and the same would happen with the incidences (since the total population does not change). This produces incidences only about of the order of 5 times larger than those in Fig.1, compatible with expectations, see SM (section I.D).

**FIG. 4.**
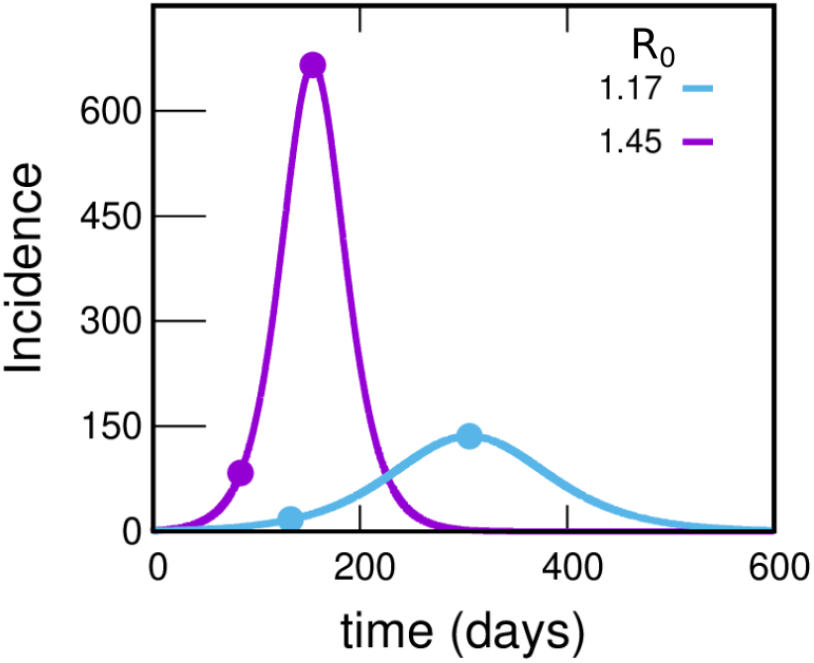
Incidences within the uniform mixing approximation for two values of *R*_0_. In each case the circles indicate the position of the maximum of the curve and another point with an incidence value 8 times lower. The violet and light-blue curves integrate to 55.0 and 27.6% of the population, respectively. Either this percentage or the time at which the peak occurs misses those observed in reported data by much more than can be reasonably expected, pointing to a failure within this simple approach. The uniform mixing approximation is achieved in our model by taking: *β*_1_ = *β*_2_ = …*β*_*k*−1_ = 0, *β*_*k*_ = *R*_0_ * *γ, f*_*C*_ = 1. These curves were obtained by solving the deterministic equations (which correspond to the limit *N* →∞) starting from an initial condition where 99.99% of the population is susceptible and 0.01% is in the infectious state *I*_1_.

To explore this idea in more realistic situations, we considered various plausible structures corresponding to the hierarchical model in Figure 3 that reproduce the central features of the dynamic evolution of the incidences observed in Figure 1. We consider a system of 300,000 inhabitants (size of a typical GBA district), starting from a state in which a fraction *f*_*C*_ of individuals participates in the dynamics of the epidemic (active) and the rest is inactive. In the initial condition *t* = *t*_ini_, all active individuals are susceptible except *I*_ini_ individuals that are in state *I*_1_. We are not concerned here with how those initially infected entered GBA; we briefly discuss this in the SM (section III.B.2). The system is allowed to evolve freely and, in order to compare with data, reported incidences are multiplied by a factor *k* and the origin of the time axes adjusted so as to obtain the best agreement in each case.

The parameters of the five scenarios discussed in the present section are presented in Table I (more scenarios are considered in section III.A of the SM). Top panels for Fig. 5a show the incidences obtained from scenarios 1 and 2. In scenario 1, the individuals interact within the family and with any other individual in the district while in scenario 2 there is a higher hierarchy of interactions that decrease in magnitude with the spatial scale (*β*_1_ *> β*_2_ *> β*_3_ ≫ *β*_4_ ≫ *β*_5_). In both cases, incidences values are obtained that are compatible with those reported with *k* = 4.5, which is within what it is expected. In scenario 1, where there are only two hierarchical levels (i.e. the scenario where the active individuals interact almost homogeneously), the incidence shows a faster fall than in scenario 2, in which the interactions between individuals from different neighbourhoods are very low.

**TABLE I.** Parameters that define the 5 scenarios considered. Number of levels (*L*), number of individuals in level 1 (*N*_1_), number of groups of size *N*_*l*−1_ individuals that make up level *l* (*n*_*l*_, *l >* 1), household contact rate (*β*_1_), average effective contact rate between individuals of the same group in level *l* (*f*_*C*_*β*_*l*_), and mean fraction of active individuals (*f*_*C*_) defining the model. Note that the number of individuals on a group in level *l* can be calculated as *N*_*l*_ = *N*_1_*n*_2_ … *n*_*l*_, *l >* 1. *I*_*ini*_ is the initial number of infected introduced at time −*t*_*ini*_. The constant *k* is not a model parameter: it is the factor we use to multiply the reported incidences so that they approach the incidence curve obtained for each scenario (see Figs. 5-7). In a similar way, *t*_*ini*_ (in days) is determined after the simulations, so that it maximises the agreement with the epidemiological data. We have explored many more scenarios, part of which are described in section III.A of the SM and Ref. 29. All contact rates are specified in units of 1/day.

| Scenario | $L$ | $N_1$ | $n_2$ | $n_3$ | $n_4$ | $n_5$ | $\beta_1$ | $f_C \beta_2$ | $f_C \beta_3$ | $f_C \beta_4$ | $f_C \beta_5$ | $f_C$ | $I_{ini}$ | $t_{ini}$ | $k$ |
| --- | --- | --- | --- | --- | --- | --- | --- | --- | --- | --- | --- | --- | --- | --- | --- |
| 1 | 2 | 4 | 75,000 | - | - | - | 0.214 | 0.10 | - | - | - | 0.25 | 150 | 20 | 4.5 |
| 2 | 5 | 4 | 5 | 100 | 10 | 15 | 0.214 | 0.10 | 0.05 | 0.00875 | 0.0005 | 0.25 | 150 | 10 | 4.5 |
| 3 | 5 | 4 | 5 | 100 | 10 | 15 | 0.214 | 0.11 | 0.055 | 0.01 | 0.0005 | 0.25 | 60 | 25 | 5.0 |
| 4 | 5 | 4 | 5 | 100 | 10 | 15 | 0.214 | 0.10 | 0.055 | 0.004 | 0.0005 | 0.50 | 150 | 25 | 10.0 |
| 5 | 5 | 4 | 5 | 100 | 10 | 15 | 0.214 | 0.13 | 0.05 | 0.002 | 0.00025 | 1.00 | 150 | 35 | 22.0 |

**FIG. 5.**
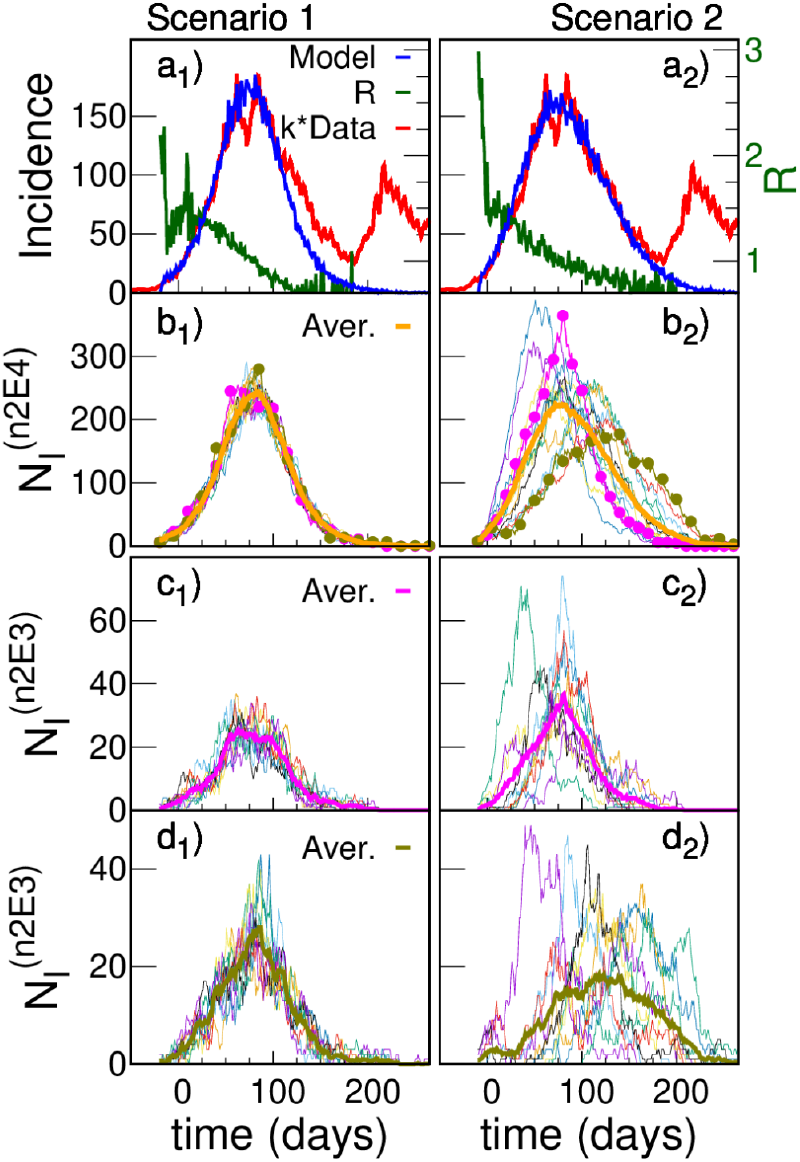
Model simulations and scaled data for two different scenarios. (a) Simulated (blue) daily cases per 100,000 inhabitants as a function of time for a total population of 300,000 for two different structures: scenario 1 (left panel) and scenario 2 (right panel); *I*_*ini*_ = 150 infected were introduced at an initial time. The green curve (right y-axes) shows that after a short transitory the simulated *R*(*t*) starts near 1.5, and decreases smoothly as the epidemic progresses. Simulations are compared with reported cases (red) in Argentina’s Greater Buenos Aires, where *t* = 0 corresponds to the 1st June 2020. In order to approach the simulation incidences, the data were multiplied by a constant factor *k* = 4.5. The idea of having only a 25% active fraction of the population leads to results comparable with reported data for a plausible value of under-reporting, and are robust regarding variations in the hierarchical structures and choice of parameters. We only show one stochastic realization of each scenario, since for these scenarios the variations between different realisations are not significant. The robustness of the idea has been tested in a greater number of hierarchies and sets of parameters, see SM (section III). (b) Infected individuals (*N*_*I*_) in each one of the 15 neighbourhoods of 20,000 inhabitants (n2E4 neighbourhoods) that make up the systems shown in (a). The thick line is the average number of infected individuals in a n2E4 neighbourhood. Note: in scenario 1, where there is no explicit separation in neighbourhoods, an arbitrary (random) aggregate of houses was made to define the corresponding neighbourhoods. (c) Infected individuals in each one of the 10 neighbourhoods of 2,000 inhabitants (n2E3 neighbourhoods) that make up two particular n2E4 neighbourhoods shown in (b). The thick line is the average number of infected individuals in each case.

The slower fall in incidences in scenario 2 can be understood by observing the evolution of those infected in the different neighbourhoods that make up the district (Fig. 5 (b) and c)). Although in both cases there are stochastic fluctuations at the different levels, in this case such fluctuations are stronger, and there is a significant lag in the beginning of the epidemic within each neighbourhood as well. In addition to this lag, there is also a marked heterogeneity in the way in which the epidemic develops in the different neighbourhoods. These differences are the origin of the slower decrease of incidence found in scenario 2 (Fig. 5 (a)). These two scenarios have been chosen just to exemplify cases of high or low coupling between neighbourhoods. We consider other cases in the SM (section III.A).

The differences in incidences that have been observed between different GBA districts (Figure S2) can be reproduced by simulating different systems with slightly different values of the parameters *β*_*l*_, *N*_*l*_ o *f*_*C*_ (which would imply attributing them to different individual behaviour in different districts). However, they can also be obtained from different simulations of a system with the same set of parameters (i.e., they are a consequence of the stochastic nature of the model, which is more evident for certain sets of parameters, as we will see in the following). Fig. 6 shows 4 realisations of a scenario 3 system, with parameters similar to scenario 2, but where initially there are *I*_ini_ = 60 infected (instead of 150). The lower number of initially infected distributed in the system in this case increases its initial heterogeneity, which translates into differences at district level that were not observed in the different realisations of scenario 2 (not shown).

**FIG. 6.**
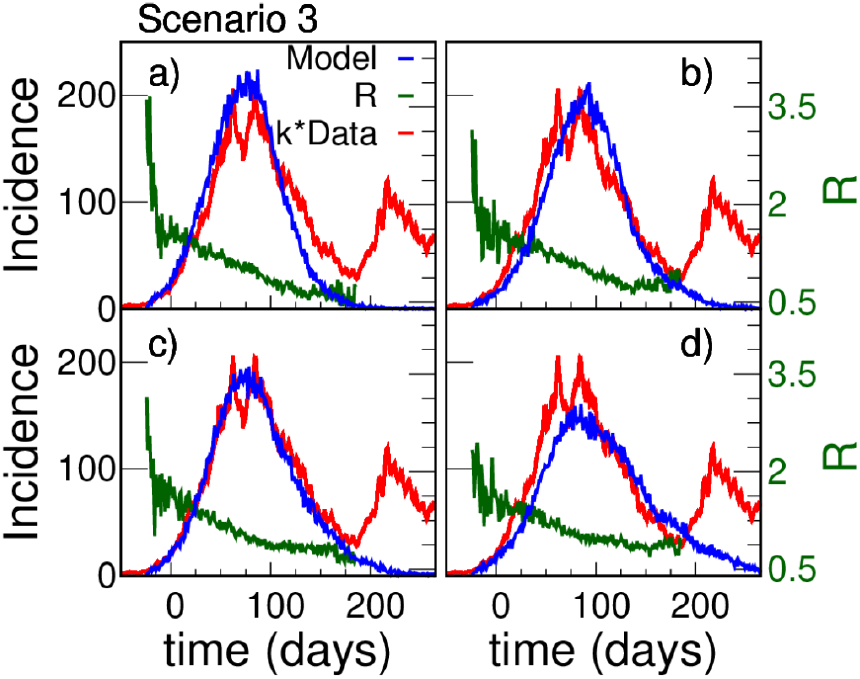
Less initially infected agents: more fluctuations. Simulated (blue) and reported (red) daily cases per 100000 inhabitants as a function of time for a total population of 300000, for the hierarchical model of scenario 3, similar to scenario 2 (shown in Fig. 5a) right panel) but for *I*_*ini*_ = 60. Panels a) to d) show different realisations of the same model with the same number of initially infected individuals that are randomly assigned in a different way into the system. We observe marked fluctuations in the behaviour, based on the stochasticity of the model and supported by its intrinsic non-homogeneous structure. Similar fluctuations have been observed on data corresponding to different districts within the GBA (Fig. S2). One of our points is to show that these fluctuations can be based on real differences on the district connective structure, or simply on the stochastic nature of the processes under way. In this case the data were multiplied by a constant factor k=5 (also compatible with a plausible under-reporting) in order to approach the simulation results.

In Fig. 7 we show the results of scenarios 4 and 5 (see Table I) which have structures similar to those of scenario 2, but also the fraction of active individuals is higher. It is seen that it is possible to reproduce the observed dynamics, but with *k* factors that are too large and incompatible with the available epidemiological information. Therefore we conclude that to obtain results compatible with the data, *f*_*C*_ must be lower than 0.50.

**FIG. 7.**
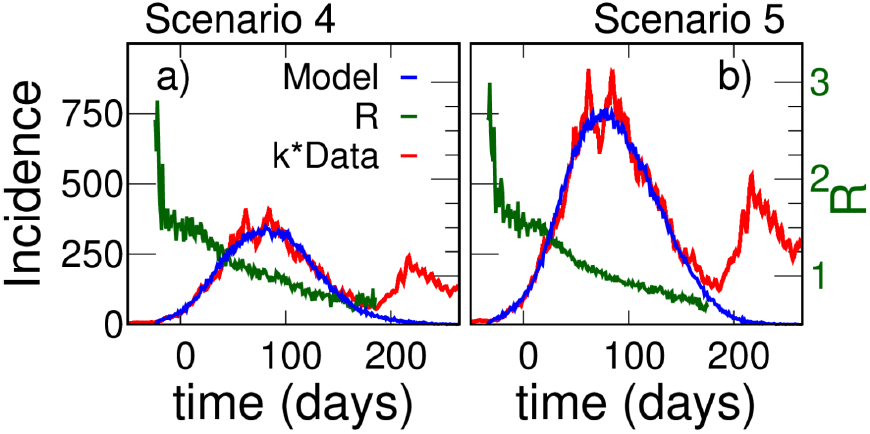
Other fractions of active individuals. Panels a) and b) correspond to scenarios 4 and 5, with *f*_*C*_ = 0.50 and *f*_*C*_ = 1.0, respectively. It is seen that the dynamics of the curve can also be reproduced as in scenario 2 (Fig. 5, right panel) but the required k-factors (10 for case 4 and 22 for case 5) are not compatible with the epidemiological information available.

This picture, in which there is a relatively small fraction of active individuals, implies a potential risk: that the majority fraction who remained inactive would join the arena –either because they stop taking care of themselves or because they get involved in activities in which they did not participate. This may have been at the bottom of what happened in Argentina at the beginning of December when there was a significant increase in cases that had its maximum near the new-year celebrations. It is possible that this rise was the result of two effects: more people becoming active, and an increase of infectious contacts. In order to illustrate the consequences of the first of these effects, two simulations (f25 and f50 in Fig. 8) were performed where additional fractions of 25 and 50% of individuals were set active by the end of November, preserving the same effective contact rates as the previous active individuals (same values of *β*_1_ and *f*_*C*_*β*_*l*_, for *l >* 1). The results are shown in Fig.8. Panel *a*_2_) shows that freeing an additional 50% of the population produces a curve compatible with the rate of rise of incidences observed in the reported data. There were many changes in the epidemiological situation beyond the end of November 2020; this accounts for the structure observed in the data shown in Fig. 8, which is beyond what we are trying to model here. The purpose of this study concerning dates after November 2020 is to show that the huge increase in cases that was observed at the end of 2020, and later in 2021 (Fig. 8) is compatible with a significant number of individuals who remained susceptible and began to interact.

**FIG. 8.**
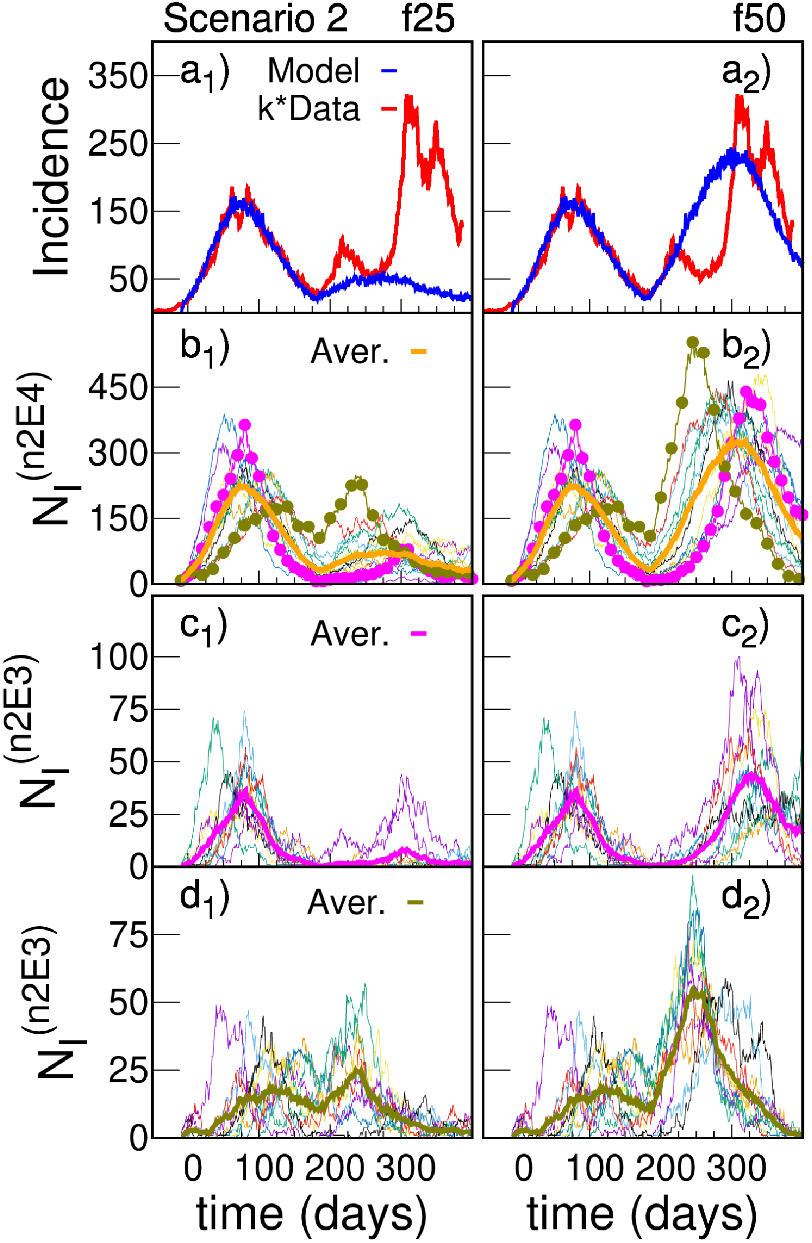
Instability: explosion of cases when the inactive become epidemiologically active. The panel structure is the same as in Fig. 5, now exploring scenario 2 beyond the end of November. Panels a) show the simulated (blue) and reported (red) daily cases per 100000 inhabitants as a function of time for a total population of 300000, for *I*_*ini*_=150 initially infected. As in Fig. 5, right panel, the simulations were performed with 75% of the population remaining inactive regarding the infection or transmission of the virus. However, at day 180 (in correspondence with end of November) this fraction was reduced by freeing (changing its state from inactive to active) a fraction of the population of 25% (f25, left panel) and 50% (f50, right panel). There is a clear correspondence with a minor “second wave”, shown as a peak in the data. Our simulations clearly signal the potential risks of this highly metastable situations (particularly in the second huge peak seen in panel a_2_), blue curve). Panels b) explore the different n2E4 neighbourhoods, and panels c) and d) what happened for different n2E3 within two n2E4 neighbourhoods.

## IV. DISCUSSION

Before drawing conclusions, some considerations regarding the hypotheses and the results must be made, given the uncertainties concerning the epidemiological data, as well as those that still exist about the characteristics of the disease.

In order to compare the results of the simulations (which provide with the number of all the “real” cases once a model and a given realisation of randomness are assumed) with the epidemiological data (which suffers from under-reporting) we have multiplied the latter by a time-independent factor *k*. Based on previous works^30–35^ we estimate the value of *k* between 3 and 6 (see SM: Sec. I.D)). A first remark is that, as we noted at the end of Sec. IIA, the factor *k* might not be constant over time, which can affect the shape of the curve of real cases in a way that is difficult to predict. At this point it is then very important to differentiate what may be considered an artificial feature, and what a solid fact. For example, it is not so straightforward to conclude from the comparison of the red and blue curves in panels a_1_) and a_2_) of Fig. 5, that Scenario 2 (highly heterogeneous) describes the spread of COVID-19 in GBA better than Scenario 1. On the other hand, given that the definition of a suspect case became more inclusive over time,^36^ and that there is no reason to think that the vigilance has been greatly relaxed, the massive drop in cases observed in Fig. 1 from September onward remains a basic feature that any reasonable proposal should try to explain.

A second remark is that the slow decline of reported cases (panels a) in Fig. 5 could be the consequence of an increase in the values of the contagion rates (which we have assumed constant) since September, rather than due to the stochastic heterogeneity among different neighbourhoods proposed within scenario 2. Although a hypothetical increase in rates could be attributed to the slight observed increase in mobility or a relaxation of preventive measures, this could have been compensated by the fact that more activities were carried out in open spaces (due to increasing temperatures) which would lower those rates.

However, there is independent epidemiological evidence that points in the direction of a heterogeneous scenario in the spread of COVID-19 in the GBA. In a seroprevalence study carried out in neighbourhoods of different districts of the GBA,^34^ a high heterogeneity was found without a clear explanation, sometimes finding very different values for seroprevalence in nearby neighbourhoods with similar characteristics. According to our simulations, stochasticity in weakly coupled neighbourhoods could account for this phenomenon. For example, looking at two of the n2E4 neighbourhoods of panel b_2_) of Fig. 5, we see that the fraction of individuals recovered up to *t* = 150 (October 18) in the neighbourhood indicated with pink circles is 18%, while it is only 11% for that with green circles. Moreover, for that same date, the fraction of individuals recovered in each of the n2E3 neighbourhoods of panel d_2_) of Fig. 5 fluctuates between 4 and 19%. Since only 25% of individuals on average can get sick (*f*_*C*_=0.25), this spread of values is markedly heterogeneous.

An important point is that the description that emerges from scenario 2 is not the consequence of a particular choice of model parameters. We check in the SM that the results are robust under changes within plausible values of these parameters (section III.A), under the consideration of the distribution of household sizes corresponding to GBA (section III.B.3), and also under the possibility of an increasing income of infections originated in CABA, where many GBA residents work (section III.B.2).

A third remark concerns natural immunity. Even though the mechanisms that generate natural immunity are not fully understood and are subject of research, several studies estimated that infection with SARS-CoV-2 provided 80-90% protection from reinfection for up to 7 months.^2,3,5^ Natural immunity would confer protection against symptomatic and also asymptomatic reinfections. Reinfection is rare but has also been reported. In order to simplify the approach, we haven’t considered this possibility here, but we explore the issue in the SM (section III). Using a deterministic model and assuming that 20% of the population does not acquire immunity after infection, we find that the impact of the effect is low: the rate of increase in incidence is basically unchanged, and the peak is reached for a value 18% higher. We therefore expect that the inclusion of this effect in the hierarchical model will not produce qualitative changes beyond a moderate increase in the value of k necessary to approximate the data.

It is important to acknowledge that there exist previous works discussing the possible reduction of the disease-induced herd immunity level as a consequence of the fact that social contacts in the population can be very inhomogeneous.^37,38^ The idea is that the better connected individuals tend to become infected (and then, recovered and immune) before the less connected ones. The latter, with a much lower potential to infect or become infected, become increasingly predominating within the active population; this naturally lowers the immunity fraction needed to stop the propagation of the disease. In a way, our assumption that only a minority fraction participates on the contagion process takes this idea of heterogeneity to an extreme. In spite of this, it is important to realise (as exemplified in Fig. 8) that in our case it is not a real herd immunity, since inactive individuals constitute a pool of susceptibles with the potential to become active.

The fourth remark regards pre-existing immunity to SARS-CoV-2. Detection of SARS-CoV-2-reactive CD4+ T cells in unexposed individuals, suggests cross-reactive T cell memory between circulating “common cold” coronaviruses and SARS-CoV-2.^39^ However, it has been argued that pre-existing SARS-CoV-2-specific T cells are unlikely to provide sterilising or herd immunity, and it is unclear whether it even affects the severity of the disease.^40,41^ So, it does not appear that pre-existing immunity affects the results of this study.

Finally, weather may play a role that is not trivial to analyse. Low temperatures are usually associated with less ventilation and more transmission. This was probably not a great influence, since the curve stops growing and starts to go down during August when it is still relatively cold in this region (mean temperatures vary from ≈ 11 in July to ≈ 13 Celsius in August^42^). On the other hand, at the beginning of the southern summer the curve was in full rise but, as mentioned, the leading effect at that time was probably from the social new-year gatherings. In any case, it cannot be ruled out that the weather may have had some influence.

## V. SUMMARY AND CONCLUSIONS

The evolution of incidences in Greater Buenos Aires presents certain characteristics shared by the 24 districts that comprise it. The incidence curve goes though a peak with a fast increase during June and July, and a somewhat slower downward slope encompassing September, October, and November. This evolution occurred under very particular conditions. The six-month period between June and the end of November 2020 was *before* the massive application of vaccines; also —and in contrast with other peaks observed during the COVID-19 pandemic in other places— the important social and hygienic measures had been taken before the fast incidence increase, and remained fixed during the curve development. Here we have shown that a coherent explanation of this evolution can be obtained if it is assumed that a minority fraction of the population participated in the contacts, so that the incidences decreased as this group of people was becoming immune. This picture is compatible with the strong outbreaks seen later, which point to the existence of a considerable pool of susceptible individuals. Our explanation implies that the evolution of the incidences in GBA would be the epidemiological manifestation of immunity conferred by SARS-CoV-2 infection.

The hypotheses assumed account for the observed behaviour without the need to fine-tune the set of parameters. This is one of the strengths of our model, which — although it could be tuned to match closely the available data— we use to explain the curve drop and to analyse a number of alternatives consistent with the several factors that may have influenced the dynamics within the epidemiological uncertainties.

Taking as true the rate of rise of reported incidences in June and July 2020, our model predicts a *basic reproduction number R*_0_ around 1.5 during epidemic spread. On the other hand, the fraction of active population *f*_*C*_ can not be accurately determined, since it depends on the actual number of cases that (due to under-reporting) can only be estimated roughly. One of our hypothesis is that individuals interact with markedly decreasing strength as one considers wider circles of contacts. This is compatible with the fact, supported by epidemiological evidence, of great heterogeneity in the transmission process in GBA. This heterogeneity, plus the intrinsically stochastic nature of the transmission process means that, even under the same conditions, results of the transmission at local levels can be very different in different districts. This constitutes a difficulty when trying to evaluate the effects of a public health policy, because it may be implemented in the same way in two places but result in a very different number of cases.

## Data Availability

Most of the data used in the present study is available online: https://datos.gob.ar/dataset/salud-covid-19-casos-registrados-republica-argentina/archivo/salud_fd657d02-a33a-498b-a91b-2ef1a68b8d16, and the rest can be requested at https://www.argentina.gob.ar/solicitar-informacion-publica Simulation software in C++ was developed for this work, and is available in the GitHub repository, https://github.com/tgrigera/COVIDm

## VI. ACKNOWLEDGEMENTS

We acknowledge financial support from the Agencia Nacional de Promoción Científica y Tecnológica (ANPCyT) through PICT 2017-2347, and the Consejo Nacional de Investigaciones Científicas y Técnicas (CONICET) through PIP 0446 and PIP 0292. G.F. wishes to aknowledge helpful discussions with colleagues from the “Red de Modelización de Enfermedades Infecciosas” (RITS-CONICET) and from the “Subcomisión de Vacunología” (Argentine Association of Microbiology).

## VII. DATA AVAILABILITY

Simulation software in C++ was developed for this work, and is available in the GitHub repository, https://github.com/tgrigera/COVIDm.

Data was provided by the Health Ministry of Argentina through the National Science Council (CONICET). Most of the data is available in the following site: https://datos.gob.ar/dataset/salud-covid-19-casos-registrados-republica-argentina/archivo/salud_fd657d02-a33a-498b-a91b-2ef1a68b8d16, and the rest can be requested at https://www.argentina.gob.ar/solicitar-informacion-publica since in Argentina data is public by law 27.275 “Derecho a la información pública”.

## Notes

### Competing Interest Statement

The authors have declared no competing interest.

### Funding Statement

This study was funded by the Agencia Nacional de Promocioacute;n through PICT 2017-2347, and the Consejo Nacional de Investigaciones Científicas y Técnicas (CONICET) through PIP 0446 and PIP 0292.

### Author Declarations

Data was provided by the Health Ministry of Argentina through the National Science Council (CONICET). Most of the data is available in the following site: https://datos.gob.ar/dataset/salud-covid-19-casos-registrados-republica-argentina/archivo/salud_fd657d02-a33a-498b-a91b-2ef1a68b8d16, and the rest can be requested at https://www.argentina.gob.ar/solicitar-informacion-publica since in Argentina data is public by law 27.275.

## References

1 Robin N Thompson, T Déirdre Hollingsworth, Valerie Isham, Daniel Arribas-Bel, Ben Ashby, Tom Britton, Peter Challenor, Lauren HK Chappell, Hannah Clapham, Nik J Cunniffe, et al., “Key questions for modelling COVID-19 exit strategies,” Proceedings of the Royal Society B 287, 20201405 (2020).

2 World Health Organization, “COVID-19 natural immunity, Scientific brief, May 10th 2021,” https://apps.who.int/iris/bitstream/handle/10665/341241/WHO-2019-nCoV-Sci-Brief-Natural-immunity-2021.1-eng.pdf, accessed: 2021-11-15.

3 Victoria Jane Hall, Sarah Foulkes, Andre Charlett, Ana Atti, Edward JM Monk, Ruth Simmons, Edgar Wellington, Michelle J Cole, Ayoub Saei, Blanche Oguti, et al., “SARS-CoV-2 infection rates of antibody-positive compared with antibody-negative health-care workers in England: a large, multicentre, prospective cohort study (SIREN),” The Lancet 397, 1459–1469 (2021).

4 Jennifer M Dan, Jose Mateus, Yu Kato, Kathryn M Hastie, Esther Dawen Yu, Caterina E Faliti, Alba Grifoni, Sydney I Ramirez, Sonya Haupt, April Frazier, et al., “Immunological memory to SARS-CoV-2 assessed for up to 8 months after infection,” Science 371 (2021).

5 Christian Holm Hansen, Daniela Michlmayr, Sophie Madeleine Gubbels, Kåre Mølbak, and Steen Ethelberg, “Assessment of protection against reinfection with SARS-CoV-2 among 4 million PCR-tested individuals in Denmark in 2020:a population-level observational study,” The Lancet 397, 1204–1212 (2021).

6 Flavia Riccardo, Marco Ajelli, Xanthi D Andrianou, Antonino Bella, Martina Del Manso, Massimo Fabiani, Stefania Bellino, Stefano Boros, Alberto Mateo Urdiales, Valentina Marziano, et al., “Epidemiological characteristics of COVID-19 cases and estimates of the reproductive numbers 1 month into the epidemic, Italy, 28 January to 31 March 2020,” Eurosurveillance 25, 2000790 (2020).

7 Google LLC, “Google COVID-19 community mobility reports,” https://www.google.com/covid19/mobility/, accessed: 2021-02-01.

8 Transport Ministry of Argentina, “Number of SUBE card users per day in AMBA,” https://datos.transporte.gob.ar/dataset/sube-cantidad-de-tarjetas-usuarios-por-dia-en-amba, (in Spanish), accessed: 2021-10-23.

9 Health Ministry of Argentina through the National Science Council (CONICET), part of that database is available in the following site: https://datos.gob.ar/dataset/salud-covid-19-casos-registrados-republica-argentina/archivo/salud_fd657d02-a33a-498b-a91b-2ef1a68b8d16, accessed: 2021-07-06.

10 Argentinian National Institute of Statistics and Censuses (INDEC), “Population estimates by sex, department and calendar year 2010 2025,” https://sitioanterior.indec.gob.ar/ftp/cuadros/poblacion/proyeccion_departamentos_10_25.pdf, in Spanish. Accessed: 2021-06-17.

11 Wikipedia, “Health measures for the COVID-19 pandemic in Argentina,” https://es.wikipedia.org/wiki/Medidas_sanitarias_por_la_pandemia_de_COVID-19_en_Argentina, (in Spanish). Accessed: 2021-08-15.

12 Heather Reese, A Danielle Iuliano, Neha N Patel, Shikha Garg, Lindsay Kim, Benjamin J Silk, Aron J Hall, Alicia Fry, and Carrie Reed, “Estimated incidence of coronavirus disease 2019 (COVID-19) illness and hospitalization, United States, February to September 2020,” Clinical Infectious Diseases 72, e1010–e1017 (2021).

13 Wu, Sean L., et al, “Substantial underestimation of SARS-CoV-2 infection in the United States,” Nature Communications 11.1 (2020): 1–10.

14 H Juliette T Unwin, Swapnil Mishra, Valerie C Bradley, Axel Gandy, Thomas A Mellan, Helen Coupland, Jonathan Ish-Horowicz, Michaela AC Vollmer, Charles Whittaker, Sarah L Filippi, et al., “State-level tracking of COVID-19 in the United States,” Nature Communications 11, 1–9 (2020).

15 Heather Kalish, Carleen Klumpp-Thomas, Sally Hunsberger, Holly Ann Baus, Michael P Fay, Nalyn Siripong, Jing Wang, Jennifer Hicks, Jennifer Mehalko, Jameson Travers, et al., “Undiagnosed SARS-CoV-2 Seropositivity During the First Six Months of the COVID-19 Pandemic in the United States,” Science Translational Medicine (2021).

16 Böhning, Dankmar, et al, “Estimating the undetected infections in the COVID-19 outbreak by harnessing capture–recapture methods,” International Journal of Infectious Diseases 97 (2020): 197–201.

17 Pedro C Hallal, Fernando P Hartwig, Bernardo L Horta, Mariângela F Silveira, Claudio J Struchiner, Luís P Vidaletti, Nelson A Neumann, Lucia C Pellanda, Odir A Dellagostin, Marcelo N Burattini, et al., “SARS-CoV-2 antibody prevalence in Brazil: results from two successive nationwide serological household surveys,” The Lancet Global Health 8, e1390–e1398 (2020).

18 In the SM (section III.B.1) we explore the consequences of assuming that only a fraction of individuals acquire immunity.

19 Keeling Matt J. and Pejman Rohani, Modeling infectious diseases in humans and animals (Princeton University Press, 2008).

20 David J Price, Freya M Shearer, Michael T Meehan, Emma McBryde, Robert Moss, Nick Golding, Eamon J Conway, Peter Dawson, Deborah Cromer, James Wood, et al., “Early analysis of the Australian COVID-19 epidemic,” Elife 9, e58785 (2020).

21 Daniel T Gillespie, “A general method for numerically simulating the stochastic time evolution of coupled chemical reactions,” Journal of Computational Physics 22, 403–434 (1976).

22 Qun Li, Xuhua Guan, Peng Wu, Xiaoye Wang, Lei Zhou, Yeqing Tong, Ruiqi Ren, Kathy SM Leung, Eric HY Lau, Jessica Y Wong, et al., “Early transmission dynamics in Wuhan, China, of novel coronavirus–infected pneumonia,” New England Journal of Medicine (2020).

23 Wei-jie Guan, Zheng-yi Ni, Yu Hu, Wen-hua Liang, Chunquan Ou, Jian-xing He, Lei Liu, Hong Shan, Chun-liang Lei, David SC Hui, et al., “Clinical characteristics of coronavirus disease 2019 in China,” New England Journal of Medicine 382, 1708–1720 (2020).

24 Muge Cevik, Matthew Tate, Ollie Lloyd, Alberto Enrico Maraolo, Jenna Schafers, and Antonia Ho, “SARS-CoV-2, SARS-CoV, and MERS-CoV viral load dynamics, duration of viral shedding, and infectiousness: a systematic review and meta-analysis,” The Lancet Microbe (2020).

25 National Center for Immunization and Respiratory Diseases (NCIRD) Division of Viral Diseases, “Ending isolation and precautions for people with COVID-19: Interim guidance,” https://www.cdc.gov/coronavirus/2019-ncov/hcp/duration-isolation.html, accessed: 2021-09-14.

26 Zachary J Madewell, Yang Yang, Ira M Longini, M Elizabeth Halloran, and Natalie E Dean, “Household transmission of SARS-CoV-2: a systematic review and meta-analysis,” JAMA Network Open 3, e2031756–e2031756 (2020).

27 Itai Dattner, Yair Goldberg, Guy Katriel, Rami Yaari, Nurit Gal, Yoav Miron, Arnona Ziv, Rivka Sheffer, Yoram Hamo, and Amit Huppert, “The role of children in the spread of COVID-19: Using household data from Bnei Brak, Israel, to estimate the relative susceptibility and infectivity of children,” PLoS Computational Biology 17, e1008559 (2021).

28 Carlos G Grijalva, Melissa A Rolfes, Yuwei Zhu, Huong Q McLean, Kayla E Hanson, Edward A Belongia, Natasha B Halasa, Ahra Kim, Carrie Reed, Alicia M Fry, et al., “Transmission of SARS-COV-2 infections in households—Tennessee and Wisconsin, April to September 2020,” Morbidity and Mortality Weekly Report 69, 1631 (2020).

29 Gabriel Fabricius, Tomas S. Grigera, Rodolfo A. Borzi, and José Caminos, “Numerical exploration of the conditions in which the acquisition of immunity can account for the observed drop in incidences in Greater Buenos Aires,” https://rits.conicet.gov.ar/download/informe_tecnico/Apendice-1.pdf, in Spanish. Accessed: 2020-12-3.

30 Ministry of Finance of Buenos Aires Province Government, “Provincial COVID-19 seroprevalence survey (in Spanish),” www.estadistica.ec.gba.gov.ar/dpe/images/INFORME_SEROPREVALENCIA.pdf, accessed: 2021-07-26.

31 Laura Muñoz, Marina Pífano, Andres Bolzán, Teresa Varela, Yamila Comes, Mariana Specogna, Leticia Ceriani, Jonatan Konfino, Nicolas Kreplak, and Enio Garcia, “Surveillance and seroprevalence: Evaluation of IgG antibodies for SARS-Cov2 by ELISA in the popular neighbourhood Villa Azul, Quilmes, Province of Buenos Aires, Argentina. Scielo Preprints 2020.” https://doi.org/10.1590/SciELOPreprints.1147, in Spanish, accessed: 2021-06-17.

32 Ministry of Finance of Buenos Aires City Government, “COVID-19 seroprevalence survey methodology and definitive results (in Spanish),” https://www.estadisticaciudad.gob.ar/eyc/wp-content/uploads/2020/12/ir_2020_1509.pdf, acceded 2020-12-17.

33 Silvana Figar, Vanina Pagotto, Lorena Luna, Julieta Salto, Magdalena Wagner Manslau, Alicia S Mistchenko, Andrea Gamarnik, Ana María Gómez Saldaño, and Fernán González Bernaldo De Quirós, “Severe acute respiratory syndrome Coronavirus 2, Seroepidemiology study in Argentinian slum,” MEDICINA (Buenos Aires) 81, 135–142 (2021).

34 A. Jait, A. Silva, A. Bolzán, Ch. Ballejo, C. Pamparana, E. Bartel, E. García, F. Irassar, M. Pifano, M. Aguirre, M. de San Martín, M. Marro, and L. Muñoz, “Seroprevalence of antibodies against SARS-CoV-2 in popular neighbourhoods of Buenos Aires Province (in Spanish),” https://www.margen.org/pandemia/textos/barrios.pdf, accessed: 2021-02-22.

35 Hernan Solari, “Estimation of detectable but undetected cases,” https://rits.conicet.gov.ar/download/informetecnico/Apendice-4.pdf, in Spanish. Accessed: 2020-12-3.

36 Solari HG, Natiello MA, “Stochastic model for COVID-19 in slums: interaction between biology and public policies,” https://doi.org/10.1017/S0950268821001746, Epidemiology and Infection 149, e206, 1–15.

37 Tom Britton, Frank Ball, and Pieter Trapman, “A mathematical model reveals the influence of population heterogeneity on herd immunity to SARS-CoV-2,” Science 369, 846–849 (2020).

38 M Gabriela M Gomes, Rodrigo M Corder, Jessica G King, Kate E Langwig, Caetano Souto-Maior, Jorge Carneiro, Guilherme Gonçlalves, Carlos Penha-Gonçalves, Marcelo U Ferreira, and Ricardo Aguas, “Individual variation in susceptibility or exposure to SARS-CoV-2 lowers the herd immunity threshold, MedRxiv (2020),” https://doi.org/10.1101/2020.04.27.20081893, accessed: 2021-12-10.

39 Alba Grifoni, Daniela Weiskopf, Sydney I Ramirez, Jose Mateus, Jennifer M Dan, Carolyn Rydyznski Moderbacher, Stephen A Rawlings, Aaron Sutherland, Lakshmanane Premkumar, Ramesh S Jadi, et al., “Targets of T cell responses to SARS-CoV-2 coronavirus in humans with COVID-19 disease and unexposed individuals,” Cell 181, 1489–1501 (2020).

40 Annika C Karlsson, Marion Humbert, and Marcus Buggert, “The known unknowns of T cell immunity to COVID-19,” Science Immunology 5, eabe8063 (2020).

41 Marc Lipsitch, Yonatan H Grad, Alessandro Sette, and Shane Crotty, “Cross-reactive memory T cells and herd immunity to SARS-CoV-2,” Nature Reviews Immunology 20, 709–713 (2020).

42 National Meteorological Service, “Climate of Argentina,” https://www.smn.gob.ar/estadisticas, (in Spanish). Accessed: 2021-12-10.

