## Supplementary Material for "Immunity acquired by a minority active fraction of the population could explain COVID-19 spread in Greater Buenos Aires (June-November 2020)"

(Dated: December 20, 2021)

### I. EPIDEMIOLOGICAL AND MOBILITY DATA

#### A. Mobility Data

To get an idea of the mobility trends in the population in the time period covered by the present study we present in Fig. S1 data taken from two different sources. In Fig.S1(a) we average the data taken from ref. 1 for the 24 districts that make up Greater Buenos Aires (GBA). The data was obtained by Google from cell phones of individuals who consented to have their location activated. In Fig. S1(b) we take the data from the SUBE (acronym for "Unique Electronic Ticket System", in Spanish). In this case the data corresponds to the Buenos Aires Metropolitan Area (AMBA, a bigger area that includes the GBA we studied here and the city of Buenos Aires or CABA) and consists of the computation of all the daily trips made by individuals in different means of transport. At the baseline the total number of transactions was around 3,500,000 for bus, 750,000 for train and 500,000 for subway. The subway runs only within CABA and not in Greater Buenos Aires. Population in AMBA is around a 50% greater than that of GBA.

Boths sets of curves indicate that, within the studied period (beginning of June - end of November) there were no sharp changes. If anything, the data suggest a gradual relaxation of the strict measures taken at mid March. In particular, in Fig. S1a) (that only includes GBA data) mobility trends show less variation between June and November than between march and June.

#### B. Incidences from reported data

In order to study the time evolution of COVID-19 epidemic in Greater Buenos Aires we analyzed the number of confirmed cases as a function of the Symptom-Onset-Date (SOD). There is a variable percentage of unreported SODs for different districts, with an average of 27.6% missing SOD along the GBA in the studied period. In order to obtain this information, we estimated it from the First-Sampling-Date (FSD) (one of the two dates is always recorded except for around 1% of the cases). To compute the incidences of Fig. 1 (GBA, see main text) and Fig. S2 (each district of GBA) we proceeded as follows:

1. We estimated the probability distribution of the difference between the FSD and the SOD (see Fig. S3).
2. We considered each confirmed case in the GBA, and when the SOD was missing we generated it from the FSD subtracting a random time weighted using the distribution of Fig. S3,
3. We repeat step 2) 100 times obtaining 100 different curves for each district corresponding to different random generations of the missing data.
4. We computed the incidences considering the population of each district according to 2010 census corrected by a multiplying factor 1.136 in order to estimate the population growth up to June 2020 (see section II.A in main text).
5. We finally smeared the data with a square window of three days.

#### C. Seroprevalence studies

There are several serological surveys carried out in the region of AMBA during 2020 in order to estimate the prevalence of SARS-CoV-2 antibodies in the population. There are two studies conducted by state agencies that cover extended areas including downtown areas and peripheral neighbourhoods. One performed in CABA between on September 8th and October 18th where 2024 samples were taken giving a prevalence of 10.1%[4] and the other carried out in AMBA (excluding CABA) during October and November where 452 positive results were obtained giving a prevalence of 11.6%.[5] In other study conducted in popular neighbourhoods of GBA between July 16th and December 1st a mean prevalence of 13.1% was obtained but with strong variations among different neighbourhoods.[6] In particular in five neighbourhoods of

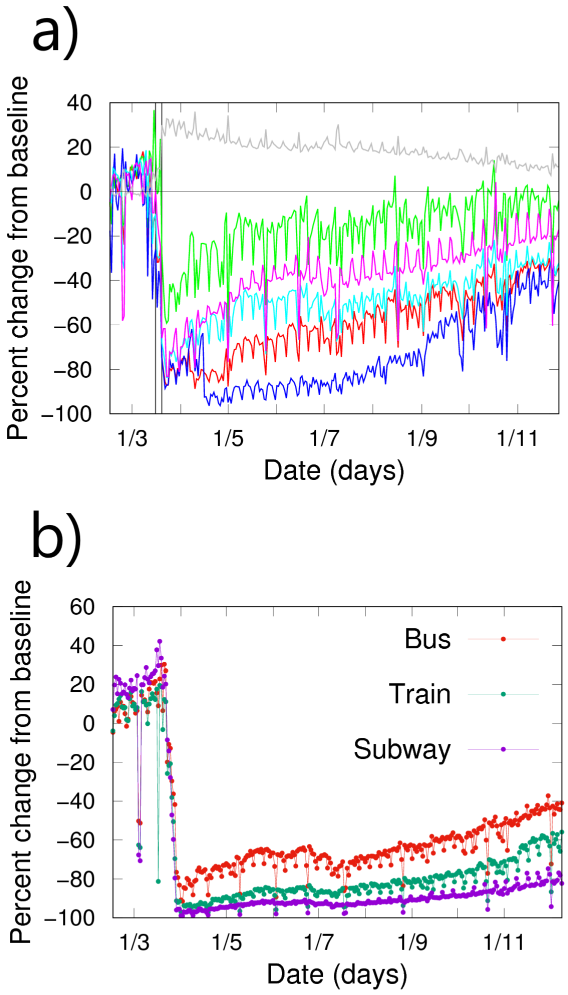


Fig. S1. **Mobility trends in 2020.** Percent changes from a baseline taken as the median value from the five-week period Jan 3rd – Feb 6th, 2020.[1] The date in the x-axis is in notation "day/month". a) Data taken from Google database for Greater Buenos Aires: 24 districts near Ciudad Autónoma de Buenos Aires (CABA). Residential (gray), grocery & pharmacies (green), workplaces (magenta), transit stations (light blue), retail & recreation (red), and parks (blue). b) Data taken from Transport Ministry (Argentina) for Buenos Aires Metropolitan Area (AMBA): 40 districts (including Greater Buenos Aires) and CABA.[2]

La Matanza district (Palito, Los Ceibos, Las Antenas, 22 de Enero, and 17 de Noviembre) prevalences of 9.3%, 10.1%, 29.1%, 23.9% and 56.7% were obtained respectively. The strong heterogeneity in the prevalence values in nearby neighbourhoods with similar characteristics was highlighted by the authors as a phenomenon to be explained.[6] On the other hand, the huge differences obtained for the prevalence in the slums Barrio Padre Mugica (CABA) in June (53.4%)[7] and Villa Azul (Quilmes, GBA) in July (14.8%)[8] has been attributed to differences in the health policy implemented in each case.[8]

#### D. Estimation of the under-reporting factor in the GBA

The incidences estimated from confirmed reported data in section I B are a lower bound to the incidences that would be obtained from the (unknown) actual cases of COVID-19. An estimation of the under-reporting factor, $f_U$, defined as the quotient between reported and actual incidences, is needed in order to compare the data with simulation results, and to have a criterion to accept or discard possible epidemiological scenarios simulated with our model. The reciprocal factor $k = 1/f_U$ accounts for the number of *actual* cases that exist for each reported one. Under-reporting may be originated in asymptomatic cases or in those with a mild symptomatology, and also in inefficiency of the detection system. The under-reporting factor may vary from place to place and in the course of the epidemic. In any case its estimation is difficult due to the impossibility of quantifying precisely the missing cases.

A rough estimation of under-reporting is usually obtained as the quotient between seroprevalence and accumulated incidence. This gives values for $k$ around 3 and 4 taking the values of prevalence mentioned in the previous section, with exception of the case of the slum Padre Mugica. There, the housing conditions led to a huge outbreak in May 2020 and a value of $k \approx 10$ was estimated.[7] A similar value ($k \approx 10.3$) was estimated in Brazil, where an extended study along the country was conducted during May and June 2020.[9] In this case, the authors of the study attributed the large value of $k$ to the controversial management of the pandemic by the national government, that restricted testing to individuals with severe symptoms during the early stages of the pandemic.

A more accurate estimation of $k$ should take into account that negative results have been obtained in serological surveys among people who had previously been diagnosed as confirmed cases of COVID-19.[5,8] In particular, in the extended study of Ref. 5 it was found that 4.8% of the sampled people had previously been diagnosed as confirmed cases, but only 42% of them were positive for the antibody test. Probably, the 11.6% of the whole sample that was positive for the antibody test is also a fraction, $f_+$, of all the cases present in the sample. If we assume that this fraction $f_+$ is approximately the same as the fraction ($f_{+D}$=0.42) that gives positive for the antibody test among confirmed (detected) cases of COVID-19, we may obtain an estimation of the total amount of the cases, $N_C$:

$$f_{+D} = 0.42 \cong f_+ = \frac{N_+}{N_C} = \frac{0.116P}{N_C} \qquad (1)$$

where $P$ is the total population, $N_+$ is the total amount of people estimated as seropositive (11.6%), and $N_C$ is the total amount of cases that can be estimated from Eq. 1 as $N_C \cong 0.276P$. This gives, if we call $N_D$ the

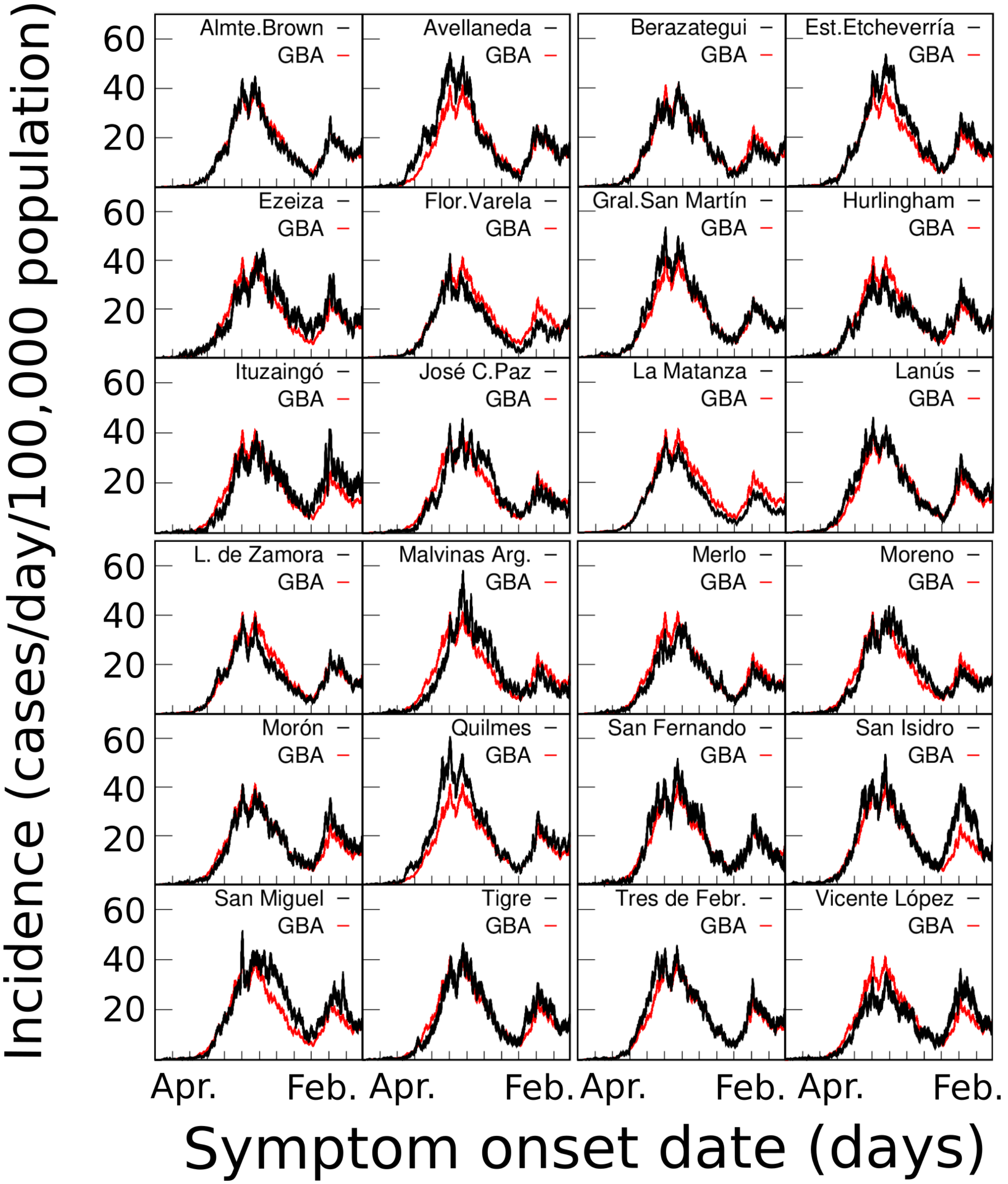


Fig. S2.

COVID-19 incidences vs time for each one of the 24 districts that make up the Greater Buenos Aires (GBA). The districts are compared with the GBA incidence as a function of the Symptom Onset Date (SOD), estimated from reported cases.[3] First and last $x-$tics stands for April 2020 and February 2021, respectively. Details of the estimation of these incidences from reported data are given in SM section I B.

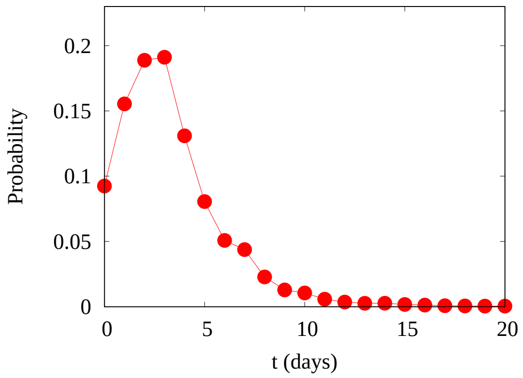


Fig. S3. **Probability that the first Sampling is taken a time $t$ after the onset of symptoms.** A total of 272,894 cases where both the SOD and the FSD are recorded in the interval from June 1st and November 30th in the GBA region were used to construct this histogram. The thin line is to guide the eye.

number of previously diagnosed cases:

$$k = \frac{N_C}{N_D} \cong \frac{0.276P}{0.048P} = 5.6 \qquad (2)$$

If this estimation is repeated taking the data from Ref. 5 but including only the districts belonging to the GBA the same estimation $k \cong 5.6$ is obtained.

In a different approach followed in Ref. 10 a lower bound estimation for $k$ is obtained by using the proportion of detected vs deceased cases in a given district. The idea was to compute this fraction for health workers (assuming that they are frequently and closely monitored and there are few missing cases), and compare it with the same proportion for the whole population. Following this approach a value of $k = 2.6$ was estimated for Buenos Aires Province.

In summary we will assume that plausible average-values for $k$ in the GBA in the studied period (June-November 2020) are between 3 and 6.

### II. MATHEMATICAL MODEL AND PARAMETERS

In this section we give some details about the contact rates in the hierarchical model described in section II.B, and some extra information about the parameters taken for the model.

#### A. Effective contact rates in the hierarchical model

In our model, each active individual $j$ at the level $l$ is associated to a given group $\nu_{l,j}$, and interacts homogeneously with all the other active individuals belonging to the same group. The rate of infection of a susceptible individual $j$ is given by expression (1) of the main text,

$$W^j_{\text{inf}} = \sum_{l=1}^{L} \beta_l \frac{N_{\nu_{l,j}}(I_1) + N_{\nu_{l,j}}(I_2)}{N_{\nu_{l,j}} - 1}$$

where $N_{\nu_{l,j}} = N_l$ is the total number of individuals in group $\nu_{l,j}$, and $N_{\nu_{l,j}}(I_1)$ and $N_{\nu_{l,j}}(I_2)$ are the number of individuals in states $I_1$ and $I_2$ respectively. However, since at a given time we consider that only a fraction $f_C$ of individuals actually participates in the infection dynamics, we define an effective contact rate, $\beta^{\text{eff}}_{l,j}$, such that the contribution to the probability of infection of a susceptible individual $j$ by the other members of group $\nu_{l,j}$ is given by

$$\beta^{\text{eff}}_{l,j} \left( \frac{N_{\nu_{l,j}}(I_1) + N_{\nu_{l,j}}(I_2)}{N^{\text{act}}_{\nu_{l,j}} - 1} \right) \qquad (3)$$

where $\beta^{\text{eff}}_{l,j}$ is the number of contacts that individual $j$ has per unit of time with other individuals of group $\nu_{l,j}$ and the factor $(N_{\nu_{l,j}}(I_1) + N_{\nu_{l,j}}(I_2))/(N^{\text{act}}_{\nu_{l,j}} - 1)$ is the probability that the contact occurs with an infected individual, since $N^{\text{act}}_{\nu_{l,j}} - 1$ is the number of active individuals in group $\nu_{l,j}$ except individual $j$. Throughout this work, *contact*, refers to *infectious contact*, a contact such that if one individual is infectious and the other susceptible, the later becomes infected.

So, comparing the term for level $l$ in 3 with Eq.(1) of main text, we obtain that $\beta^{\text{eff}}_{l,j}$, is related with the contact parameters $\beta_l$ by the expression:

$$\beta^{\text{eff}}_{l,j} = \beta_l \frac{N^{act}_{\nu_{l,j}} - 1}{N_l - 1}. \qquad (4)$$

Since, on average, the number of active individuals is $f_C N_l$, for $N_l >> 1$, $\beta^{\text{eff}}_{l,j} \approx f_C \beta_l$. But there will be fluctuations: in the groups where the active fraction of individuals is higher, so will be the effective contact rate. In summary, there are two types of heterogeneity in the contacts that the model accounts for: on one hand, there is a sharp distinction between an average fraction $f_C$ of individuals who participate in the contacts and another $1 - f_C$ that, due to the care or isolation, does not. The individuals who participate in level $l$ contacts, they do so with an average rate $\langle \beta^{\text{eff}}_l \rangle \approx f_C \beta_l$ . On the other hand, within different groups of the same level, $N^{\text{act}}$ fluctuates, and hence the effective rate $\beta^{\text{eff}}_l$.

#### B. Parametrization of the model

We have proceeded as follows: first we have chosen the values for $\sigma_1$, $\sigma_2$, $\gamma_1$, $\gamma_2$, $f_C$, $N_i$ and $I_{ini}$. Then, the choice of the contact rate parameters, $\beta_i$, is conditioned to reproduce the rate of rise observed in the data for GBA incidence. The parameter $t_{ini}$ is determined after the calculations have been carried out and defines what instant of the simulation is considered as June 1st, discarding an initial equilibration time.

**Latency and contagion times**

In our model the state of each individual is a different stochastic variable, so each individual can remain in a latency or infective state for a different time, given by a probability distribution. The choice of two latency states, $E_1$ and $E_2$, with constant rates, $\sigma_1 = \sigma_2$ (Fig. 2 and Table 1, main text), implies that once infected, the probability that the individual will become infective after a time $t$ is given by the probability distribution of Fig. S4. A very similar distribution has been obtained in Ref. 11 (Fig. 3A) from clinical data of infector-infectee pairs in South-Korea. The distribution of Fig. S4 also resembles the typical incubation period distribution[12] assuming contagion begins one or two days before symptoms onset. We have taken $\sigma_1 = \sigma_2$=2/(3 days) which gives a mean latency time $< t >=$ 3 days, compatible with an incubation period of 4-5 days.[12,13] In section III we check that taking parameters compatible with larger incubation periods (that also have been reported) does not affect the conclusions of the work.[14]

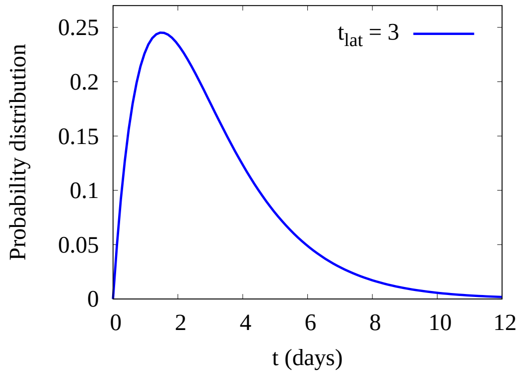


Fig. S4. **Latency time distribution.** Probability that an individual becomes infectious in the time interval $(t, t + dt)$ since the infection, given by the model for $\sigma_1 = \sigma_2 = 2/t_{lat}$. It can be computed analytically by $p(t) = 4t/t_{lat}^2 \exp(-2t/t_{lat})$.

States $I_1$ and $I_2$ are infectious states. When an individual is in any of these states, they can infect others. Therefore, the time that an individual remains in the infectious states of the model does not depend only on the clinic but on social factors. In Fig. S5 the red curve represents the probability that an individual is able to infect other people assuming that they are contagious from the day before symptoms onset until the first sample is taken, when probably isolation is recommended by health authorities. Since probably this advice is not taken until the result of the PCR test, this probability should be taken as a lower bound. Otherwise, the model accounts for all cases and not only the reported ones. Mild symptomatic or asymptomatic individuals probably infect others for a longer time being undetected although infectivity probably decreases over time. In fact, since the beginning of the pandemic there has been a lot of discussion about the duration of contagion in individuals with different symptoms.[12,15–19] But there is some consensus that contagion occurs basically during the first week of the onset of symptoms and that it is very rare for it to occur beyond 10 days.[18,19] In summary we perform most of calculations with parameters corresponding to an average infectivity time of 7 days (blue curve, Fig. S5) and take 5 and 10 days as lower and upper bounds.

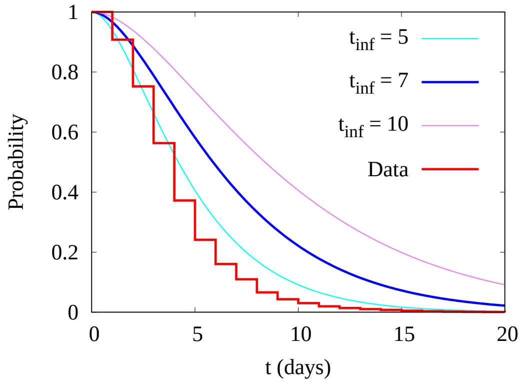


Fig. S5. **Estimations of the probability that an individual is contagious during a time $t$.** The stepped (red) curve is computed from the data of Fig. S3 assuming that an individual is contagious from the day before symptoms onset until the first sample is taken. The other curves are obtained from the model for $\gamma_1 = \gamma_2 = 2/t_{\rm inf}$ for different values of $t_{\rm inf}$ and are given by $(1 + 2t/t_{\rm inf}) \exp(-2t/t_{\rm inf})$.

**Size and structure of the system: $N_i$.**

The system to be simulated is a typical GBA-district of 300,000 inhabitants that, in most scenarios considered here, is composed by 15 neighbourhoods of 20,000 inhabitants (n2E4 neighbourhoods) that in turn are made up of 10 neighbourhoods of 2,000 individuals (n2E3 neighbourhoods). Several internal structures for the n2E3 neighbourhoods were considered. All the houses were taken of the same size ($N_1 = 4$) in most of calculations, but in section III we check that taking a distribution of house-sizes corresponding to the GBA does not modify the main results of the work.

**Initial number of infected individuals introduced in the system, $I_{ini}$**

At the beginning of the simulation $I_{ini}$ infected individuals are randomly introduced among the different neighbourhoods of the system (in section III B 2 the origin of these infected individuals is discussed). For most simulations performed we took $I_{ini}$ around 150 which leads to an average of 10 and 1 initial infecteds per n2E4 and n2E3 neighbourhoods respectively. This is consistent with the fact that, at the beginning of the studied period, the disease was quite widespread within each GBA district. However, due to fluctuations and to the fact that there is a probability that an initial infected individual recovers before infecting anybody, there will be a variable number of n2E3 neighbourhoods without infecteds at the beginning of the epidemic spread. So, taking low values for the probability contact rates among individuals from different n2E3 and n2E4 neighbourhoods lead, in general, to an heterogeneous spread in the district.

**Contact rate parameters, $\beta_i$**

These parameters, together with the $N_i$, define the contact structure of the system. For the determination of the $\beta_i$ in each structure considered we proceeded as follows: we first make a choice for $\beta_1$. Then, we determine the other $\beta_i$ regarding that (i) they fulfil the condition $\beta_1 > \beta_2 > \beta_3 >> \beta_4 >> \beta_5$ that we think was valid for the region and time studied and leads to poorly connected localities and neighborhoods, and (ii) the rate of rise and fall of the incidence of Fig.1 is well reproduced.

We also explore $\beta_3 \gtrsim \beta_4 \gtrsim \beta_5$ but in this case the structure is washed out, the system behaves as an homogeneous mixture, and it is not possible to reproduce the slow fall of the incidence curve.

*Determination of the contact rate at home, $\beta_1$*: It is the number of contacts that an individual has per unit of time with other household members. In principle, it could be estimated from the household Secondary Attack Rate (SAR), but there are not SAR measures in Argentina and the values estimated from different studies in other countries covers a very broad range.[20] The large dispersion in the reported SAR values is due to many factors beyond the characteristics of the epidemic spread, such as the type of study and the conditions of its execution. Low values of SAR are usually obtained from studies carried out within the framework of programs where the index case is isolated from other household members. We used values for $\beta_1$ corresponding to large SAR values since we are assuming that the fraction that participate of the contagion process do so without adequate hygienic measures. On the other hand, contact tracing, which may have had an impact on reducing SAR was particularly difficult to implement in the GBA region[21] and, in any case, it only affected the reported cases that are a minority fraction of the actual cases in the studied region. For most of our simulations we took $\beta_1 = 1.5\gamma \sim 0.2141$/day. For a household of size 4, using our model with $\beta_l$=0 for $l > 1$, we obtained that the probability that a susceptible individual in the household would become infected was 51%. To compute this probability we include all infections (not only secondary ones). If we restrict the time window for contagion to seven days (as in the study of Ref. 22) the probability of becoming infected reduces to 27%. These numbers seem plausible when analysing the results obtained in the study of 637 households in Bnei Brak (Israel)[23] and 101 households in Tennessee and Wisconsin (USA),[22] and the conditions in which they were made. In sensitivity analyses in section III we also perform calculations with $\beta_1 = \gamma \sim 0.1431$/day and $\beta_1 = 2\gamma \sim 0.2861$/day, which gives values of 36% and 62% respectively for the probability of becoming infected and 19% and 35% for the corresponding ones in a reduced time window of 7 days.

It is important to highlight that the parameters involved in the model are not independent. There are different set of parameters that produce equivalent descriptions of the system as is noted in section III.

### III. ROBUSTNESS OF THE RESULTS AND CONCLUSIONS

We here check the robustness of our results under changes in the model parameters and model assumptions.

#### A. Changes in the model parameters

In Table I we summarise the relevant information of the cases studied . All the scenarios correspond to a system of 300,000 inhabitants. Scenarios 1 to 5 are the ones presented in Table 2 (main text) where we observed (i) the effect of changing the structure from a set of well mixed houses to a more complex hierarchical one ( scenarios 1 and 2, respectively), that impacts in how the incidences fall after the peak, (ii) the effect of considering a high degree of heterogeneity at the beginning of the spread (low $I_{ini}$, scenario 3), that gives fluctuations in the obtained incidences comparable with those observed for different districts of GBA, and (iii) the effect of changing the fraction, $f_C$, of individuals that participate of the contagion process (scenarios 4 and 5) that defines the predicted value of the under-reporting factor ($1/k$). We concluded that $f_C$ should be lower than 0.5 in order that the model does not give incidence values unjustifiably much larger than those estimated from epidemiological data (see section I D); this excludes scenario 5 from the possible cases. The analysis also pointed to scenarios 2 and 3 as the more plausible ones. The purpose of the additional scenarios presented here (6 to 14) is to show that this "plausibility" is not restricted to a particular combination of parameters, but there is a broad spectrum of them. Some features are kept fixed to simplify the analysis of the results: all scenarios correspond to 4-members household living in neighbourhoods of 2,000 individuals (n2E3 neighbourhoods) which are connected with each other forming neighbourhoods of 20,000 individuals (n2E4 neighbourhoods) that are very weakly connected.

Scenarios 6 and 7 have the simplest possible structure inside the n2E3 neighbourhoods (i.e., each n2E3 group consists of 500 families). The main difference between both cases is that in scenario 7 the n2E3 neighbourhoods

TABLE S1. Parameters that define the 14 cases or *scenarios* considered here. Number of levels ($L$), number of individuals in level 1 ($N_1$), number of groups of size $N_{l-1}$ individuals that make up level $l$ ($n_l$, $l > 1$), household contact rate, in units of $\gamma$ ($\beta_1/\gamma$), average effective contact rate between individuals of the same group in level $l$ in units of $\gamma$ ($f_C\beta_l/\gamma$), mean fraction of active individuals ($f_C$), mean latency time, in days ($t_{lat}$), and mean infectivity time, in days ($t_{inf}$) defining the model. Note that the number of individuals on a group in level $l$ can be calculated as $N_l = N_1 n_2 \dots n_l$, $l > 1$. $I_{ini}$ is the initial number of infected introduced at time $-t_{ini}$. The constant $k$ is not a model parameter: it is the factor we use to multiply the reported incidences so that they approach the incidence curve obtained for each scenario. In a similar way, $t_{ini}$ (in days) is determined after the simulations, so that it maximises the agreement with the epidemiological data.

| Scenario | $L$ | $N_1$ | $n_2$ | $n_3$ | $n_4$ | $n_5$ | $\beta_1/\gamma$ | $f_C\beta_2/\gamma$ | $f_C\beta_3/\gamma$ | $f_C\beta_4/\gamma$ | $f_C\beta_5/\gamma$ | $f_C$ | $t_{lat}$ | $t_{inf}$ | $I_{ini}$ | $t_{eq}$ | $k$ |
|---|---|---|---|---|---|---|---|---|---|---|---|---|---|---|---|---|---|
| 1 | 2 | 4 | 75,000 | - | - | - | 1.5 | 0.70 | - | - | - | 0.25 | 3.0 | 7.0 | 150 | 20 | 4.5 |
| 2 | 5 | 4 | 5 | 100 | 10 | 15 | 1.5 | 0.7000 | 0.3500 | 0.0612 | 0.0035 | 0.25 | 3.0 | 7.0 | 150 | 10 | 4.5 |
| 3 | 5 | 4 | 5 | 100 | 10 | 15 | 1.5 | 0.7700 | 0.3850 | 0.0700 | 0.0035 | 0.25 | 3.0 | 7.0 | 60 | 25 | 5.0 |
| 4 | 5 | 4 | 5 | 100 | 10 | 15 | 1.5 | 0.7000 | 0.3850 | 0.0280 | 0.0035 | 0.50 | 3.0 | 7.0 | 150 | 25 | 10.0 |
| 5 | 5 | 4 | 5 | 100 | 10 | 15 | 1.5 | 0.9100 | 0.350 | 0.0140 | 0.0017 | 1.00 | 3.0 | 7.0 | 150 | 35 | 22.0 |
| 6 | 4 | 4 | 500 | 10 | 15 | - | 1.5 | 0.5950 | 0.1400 | 0.0035 | - | 0.25 | 3.0 | 7.0 | 150 | 21 | 4.7 |
| 7 | 4 | 4 | 500 | 10 | 15 | - | 1.5 | 0.6738 | 0.0612 | 0.0035 | - | 0.25 | 3.0 | 7.0 | 150 | 21 | 4.0 |
| 8 | 5 | 4 | 10 | 50 | 10 | 15 | 1.5 | 0.5250 | 0.3500 | 0.0612 | 0.0035 | 0.25 | 3.0 | 7.0 | 150 | 15 | 4.7 |
| 9 | 5 | 4 | 50 | 10 | 10 | 15 | 1.5 | 0.4375 | 0.3150 | 0.0612 | 0.0035 | 0.25 | 3.0 | 7.0 | 150 | 15 | 4.7 |
| 10 | 5 | 4 | 5 | 100 | 10 | 15 | 2.0 | 0.7000 | 0.3062 | 0.0612 | 0.0035 | 0.25 | 3.0 | 7.0 | 150 | 10 | 4.5 |
| 11 | 5 | 4 | 5 | 100 | 10 | 15 | 1.0 | 0.7000 | 0.3850 | 0.0612 | 0.0035 | 0.25 | 3.0 | 7.0 | 180 | 10 | 4.5 |
| 12 | 5 | 4 | 5 | 100 | 10 | 15 | 1.5 | 0.7875 | 0.4375 | 0.0525 | 0.0026 | 0.25 | 4.5 | 7.0 | 150 | 15 | 5.0 |
| 13 | 5 | 4 | 5 | 100 | 10 | 15 | 1.5 | 1.0000 | 0.4000 | 0.0400 | 0.0040 | 0.25 | 3.0 | 10.0 | 150 | 15 | 5.0 |
| 14 | 5 | 4 | 5 | 100 | 10 | 15 | 1.5 | 0.5625 | 0.3250 | 0.0612 | 0.0035 | 0.25 | 3.0 | 5.0 | 150 | 7 | 4.0 |

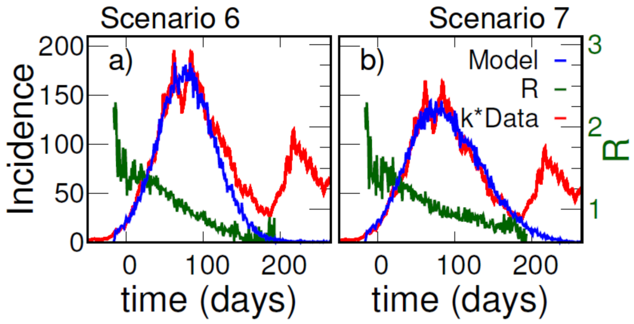


Fig. S6. **I**ncidences as a function of time for two scenarios. Simulated (blue) and reported (red) daily cases per 100,000 inhabitants as a function of time for the hierarchical structure of scenario 6 (a) and scenario 7 (b), see Table S1. In both cases, the structure is the same, and individuals have the same average number of contacts with their neighbours inside the n2E4 neighbourhoods ($\beta_2 + \beta_3$). However, the n2E3 neighbourhoods are more weakly connected (lower $\beta_3$) for scenario 7 than for scenarios 6. Simulations are compared with reported cases (red) in Argentina's Greater Buenos Aires, where t=0 corresponds to the 1st June 2020. In order to approach the simulation incidences, the data were multiplied by a constant factor $k = 4.7$ for scenario 6, and k=4 for scenario 7. The green curve (right y-axes) shows the simulated $R(t)$ computed with Ec.(2) (main text).

are more weakly connected than in scenario 6. This causes the incidence of case 6 to fall faster after the peak than that of scenario 7 (Fig. S6), as we observed for scenarios 1 and 2. However, here it is evident that the difference is related exclusively to the ratio of contact rates within n2E3 neighbourhoods and between different n2E3 neighbourhoods ($\beta_3/\beta_2$). We checked that by changing the relationship $\beta_3$ vs $\beta_2$, any intermediate behaviour for the fall of the incidences can be obtained. In scenarios 2, 8, and 9 a greater level of complexity is added with respect to case 7: the n2E3 neighbourhoods have an internal structure beyond the house. Houses are connected in groups of 5 (case 2), 10 (case 8) or 50 (case 9). For scenarios 8 and 9, incidences (not shown) present an agreement as good as for scenario 2 (panel $a_2$) of Fig. 5, main text). For this to be possible, it was necessary to decrease the values of $\beta_2$ (and also $\beta_3$ in scenario 9), that is to say that the greater availability of susceptible individuals in the second level for these cases can be offset, basically, by a decrease of the corresponding contact rate.

In the following scenarios (10 to 14) we kept fixed the structure of case 2 and study the effect of changing a single parameter in each case. In scenarios 10 (11) we check the effect of considering a greater (lower) value for $\beta_1$. In scenario 10, the same incidence behaviour than in case 2 was reproduced by lowering $\beta_3$. In scenario 11, to achieve the rate of rise observed in the data, it would have been necessary to raise the other $\beta$-values. But, increasing $\beta_2$ and $\beta_3$ does not seem very plausible, as gives $\beta_1 \sim \beta_2 \sim \beta_3$, and increasing $\beta_4$ or $\beta_5$ leads to a more rapid fall in the incidence as was observed in cases 1 or 6. In this case, it was possible to reproduce the observed incidence behaviour (for a reasonable set of contact rates) increasing the value of $I_{ini}$.

In scenario 12, $t_{lat}$ was increased by 50%. As is well known, an increase in the latency time decreases the growth rate of the epidemic.[24] Therefore, to obtain the expected growth rate it was necessary to raise the contact rates, which gave a higher $R(0) \sim 1.6$.

In scenario 13 (14) an increase (decrease) of $t_{inf}$ is considered. To reproduce the observed epidemic growth rate it was necessary to increase (decrease) the contact rates relative to the recovery rate, $\gamma$, which gives higher (lower) values of $R(0) \sim 1.65$ (1.35).

It is important to note that beyond the fact that we think the more plausible values for $\beta_1$, $t_{lat}$ and $t_{inf}$, are $1.5\gamma$, 3 days, and 7 days respectively, we have proven that a different value in each one of these parameters does not change the possibility of reproducing the observed features of epidemiological data by modifying other parameters within an equally plausible range.

The time evolution of incidences for scenarios 8 to 14 are not shown, as the behaviour is quite similar to those of scenarios 2 and 7, nearly approaching the one of GBA incidence. In fact, the objective of these simulations was to prove that this could be obtained for a variety of cases. Other result obtained from the studied scenarios concerns the value of $R(0)$, that is a good approximation to the $R_0$ of the system. We observed that in scenarios 1 to 12 it fluctuates around 1.5 within an approximately range $1.3 - 1.7$. However, fluctuations are more related to stochasticity (at the beginning of the epidemic spread) than to the different structures and parameters explored.

We have also performed simulations for $150 < I_{ini} < 300$ finding that for $I_{ini}$ up to 250 a reasonable agreement can be obtained with the epidemiological data by changing the beta within a plausible range. For $I_{ini}$ values greater than 250 it is no longer possible to approximate the epidemiological data within a reasonable range of the model parameters.

The results shown test different hierarchical structures for fixed system size of 300,000 individuals. We have also checked the robustness of our conclusions for other structures at the same size, and for some structures at system sizes of 500,000 and 1,500,000 people (not shown).

### B. Validation of some model assumptions

#### 1. *Immunity acquisition assumption*

We have assumed that, after infected, individuals acquire lasting immunity that protects them against re-infection and disease during the period of our study (6 months). Even there is enough evidence that infection with SARS-CoV-2 provided 80-90% protection from re-infection up to 7 months,[25] protection is not 100%, and some individuals may reinfect and spread this infection.

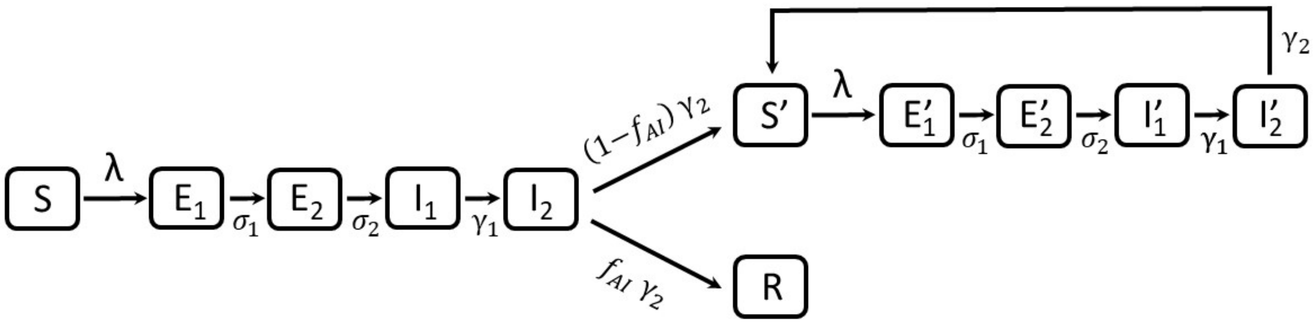


Fig. S7. **Deterministic model where only a fraction $f_{AI}$ of the infected individuals acquire lasting immunity.** When $f_{AI}$=1, the model reduces to the one used to compute the curves of Fig. 4 (main text): individuals in the susceptible (S) class are infected at a rate $\lambda$ entering a chain of exposed ($E_i$) classes where they are not infectious. They then move along the chain to the infectious classes ($I_i$) and finally recover, entering the immune class R. However, when $f_{AI} < 1$, a fraction $1 - f_{AI}$ of individuals who were infected for the first time, enter an S' class (when leaving $I_2$ class) where they may become re-infected at the same rate $\lambda$ of susceptible individuals in S class. Re-infected individuals do not acquire immunity, they perform a closed loop where they may be re-infected any number of times. $\sigma_1 = \sigma_2 = 2/t_{lat}$, $\gamma_1 = \gamma_2 = 2/t_{inf}$. The force of infection is the only time dependent rate: $\lambda = \beta(i_1 + i_2 + i'_1 + i'_2)$, where $i_j$ and $i'_j$ are the fraction of individuals in classes $I_j$ and $I'_j$ respectively ($j = 1, 2$).

To estimate how this could affect our results we have made an estimation of the order of magnitude of the effect using the deterministic model outlined in Fig. S7. The model assumes that once infected for the first time, a fraction $f_{AI}$ of the population acquires immunity while a complementary fraction may be re-infected any number of times. It is assumed that when re-infected, individuals behave and experience the disease in the same way as subjects infected for the first time. The dynamic of the model is described by the following set of differential equations:

$$
\begin{aligned}
\frac{ds}{dt} &= -\lambda s \\
\frac{de_1}{dt} &= \lambda s - \sigma_1 e_1, \qquad \frac{de_2}{dt} = \sigma_1 e_1 - \sigma_2 e_2 \\
\frac{di_1}{dt} &= \sigma_2 e_2 - \gamma_1 i_1, \qquad \frac{di_2}{dt} = \gamma_1 i_1 - \gamma_2 i_2 \\
\frac{dr}{dt} &= f_{AI} \gamma_2 i_2 \\
\frac{ds'}{dt} &= (1 - f_{AI}) \gamma_2 i_2 - \lambda s' + \gamma_2 i'_2 \\
\frac{de'_1}{dt} &= \lambda s' - \sigma_1 e'_1, \qquad \frac{de'_2}{dt} = \sigma_1 e'_1 - \sigma_2 e'_2 \\
\frac{di'_1}{dt} &= \sigma_2 e'_2 - \gamma_1 i'_1, \qquad \frac{di'_2}{dt} = \gamma_1 i'_1 - \gamma_2 i'_2
\end{aligned}
$$

where $s, e_j, i_j, r, s', e'_j$, and $i'_j$ are the fraction of individuals in the classes S, $E_j$, $I_j$, R, S', $E'_j$, and $I'_j$ respectively. The incidence is computed by: $I_{nc}(t) = \lambda(s + s')$. In Fig. S8(a) it could be seen that the magnitude of the effect is small. Even when there is a fraction of individuals that could be infected many times, it does not happen for these model parameters. Only a 43% of individuals that enter $S'$ class lead to new infections. In Fig.S8 (b) it is shown that the rate of rise of incidence is almost the same for both values of $f_{AI}$ while the curve falls down a bit slower for $f_{AI} = 0.80$. It can also be seen from the figure, that the effect could be partially absorbed by considering a lower fraction of active individuals. We mean that if $f_{AI} = 0.80$ is taken, but $f_C$=0.21, a similar description is obtained than when $f_C$=0.25 and the possibility of re-infection is neglected. We therefore do not expect the results of our study to be qualitatively modified by the inclusion of re-infections in the hierarchical model.

#### 2. *Consideration of a growing influx of infections from CABA*

We analyse here the implications and plausibility of this assumption. We discuss first the probable origin of the infected individuals originally introduced into the system. The spread of COVID-19 begins in the GBA in March from the entry of cases imported from abroad and had a very slow development at the beginning due to the severe restrictions imposed. Two factors probably contributed to the increase in the circulation of the disease observed in early June: 1) the increase in mobility that was continuous and sustained from March to June and facilitated the spread of cases that already existed in the GBA; 2) the entry of infections from CABA. It is likely that a fraction of individuals had brought the infection from CABA to GBA, since many people from the GBA districts commute to work in the city of Buenos Aires; on the other hand, it is very difficult to estimate the magnitude of this effect during the period of restrictions, and to know if it played a relevant role. This effect of im-

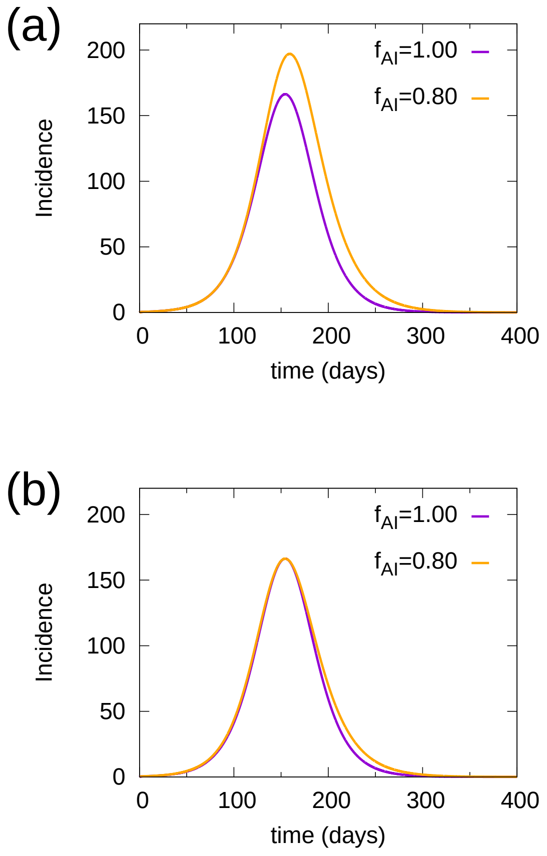


Fig. S8. **Incidences vs time predicted by the deterministic model of Fig. S7**. Incidences (in daily cases per 100,000 population) where obtained for $R_0$=1.45, $t_{inf}$=7 days, $t_{lat}$=3 days, and two different values of the fraction of the population that acquires immunity ($f_{AI}$). (a) Time evolution of the system from an initial condition (t=0) with $s$=0.9999, and $i_1$=0.0001 for $f_{AI}$=1 (violet) and $f_{AI}$=0.80 (orange). Incidences are multiplied by a factor $f_C$= 0.25, assuming the model describes the spread among a fraction $f_C$ of active population. The violet curve is the same than in Fig. 4 (main text) multiplied by $f_C$. (b) The same than (a), but the orange curve has been multiplied by a factor $f_C$=0.21 and has been shifted 5 days leftwards, so that both curves reach the maximum at the same time.

ported infections, if any, probably increased during the month of May since incidences in CABA grew strongly since 1st May (see Fig. S9) and could have dragged the growth seen in GBA from June. If this was the case, it is logical to assume that the effect continued during the following months.

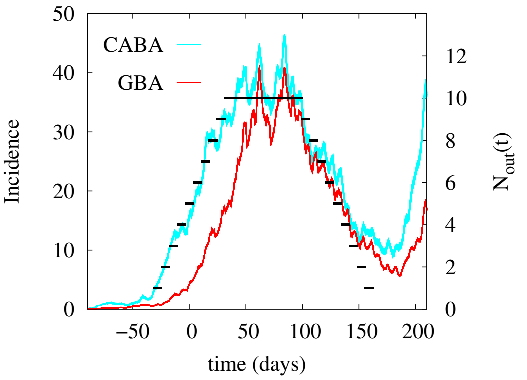


Fig. S9. **Inward flow of infected individuals from CABA**. CABA (cyan) and GBA (red) incidences vs time. A flow of infected individuals introduced in the system, $N_{out}(t)$ (black, right y-axis), is used to simulate a hypothetical entry of infections from CABA to a typical GBA-district. Incidences (in cases per day per 100,000 inhabitants) are plotted as a function of the symptom onset day, where $time = 0$ corresponds to 1st June. The steep slope of CABA incidences starts around 1st May, a month before than in GBA.

To estimate the impact of this effect (which was not considered in the previous results) we have carried out simulations in which, instead of starting with a number of initial infected, $I_{ini}$, we daily change the state of a number $N_{out}$ of individuals from S to I, assuming they have cought the infection outside the district. We take this number proportional to the incidence in CABA as is schematized in Fig. S9, assuming that the number of people who travel daily to CABA remained approximately constant over time, and that the probability of them becoming infected is proportional to the circulation of the disease in CABA. The constant of proportionality taken is such that by June 1, 90 infected had entered; at the end of June, 300, and the total number of infections entering from CABA during the whole period analised here added to 1,310. We first performed a simulation taking the parameters of scenario 2; the results are shown in Fig. S10(a). The introduction of an increasing number of infections from outside the district increases the value of the incidence at the peak in a factor 1.33 with respect to the original scenario 2, and the curve drops sharply after the peak (heterogeneous spread is lost). We then repeat the simulation in identical conditions but taking $\beta_4 = \beta_5 = 0$. In this case the incidence evolution is very similar to that of scenario 2 (See Fig. S10(b)). We conclude that the considered entry of infections from outside the district plays the same role that the contacts between individuals from different neighbourhoods in scenario 2. From our analysis, we cannot conclude which picture is closer to what really happened.

In summary, if we assume that the initial infected individuals, $I_{ini}$, considered in the simulations presented in the main text, originate in infections caught in CABA basically in May, the additional assumption that this income of infections continues in an increasing rate does not change the conclusions of our work.

On the other hand, the infections caught outside the district, $N_{out}(t)$, are probably not randomly distributed in the system, as considered. Probably individuals from some neighborhoods were more likely to work in CABA. If this was the case, we do not expect the results of the simulations shown in Fig.S10(a) to differ so much with the ones of scenario 2. Probably renormalizing all the $\beta_i$ would be enough to account for the effect.

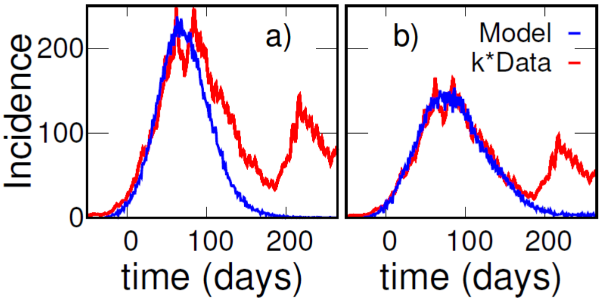


Fig. S10. **Scenarios with an inward flow of infected individuals from CABA.** Simulated (blue) and reported (red) daily cases per 100,000 inhabitants as a function of time for a non-isolated district. Differently from previous cases, we start with no infected individuals. At time $t$, the epidemiological state of a number $N_{out}(t)$ of randomly chosen *active* individuals was switched from $S$ to $I_1$. $N_{out}(t)$ is the function plotted in Fig. S9. Panel (a) shows the incidence obtained for parameters corresponding to scenario 2 (except $I_{ini}$), where the data were multiplied by a factor $k$=6 in this case. We used the same parameters in panel (b), excluding contacts between individuals from different neighbourhoods ($\beta_4 = \beta_5 = 0$). In this case, the data were multiplied by $k = 4$ (very similar to the $k$=4.5 used in scenario 2).

#### 3. *Consideration of GBA household size distribution*

In order to verify that having taken a fixed size equal to 4 for the households does not modify the conclusions of the work, we carried out simulations for a structure like the one in scenario 2, but taking a distribution of households like the one reported for the GBA[26] (see table S2). The allocation of households made up of active and inactive individuals was carried out randomly taking into account that the resulting distributions for the entire district respect the proportions of the table S2). However,

TABLE S2. Household size distribution taken for the simulations discussed in this section. The distribution corresponds to that reported in the 2010 census for Greater Buenos Aires[26] except that here we assign a size equal to 6 to the larger households.

| Household size | 1 | 2 | 3 | 4 | 5 | 6 |
|---|---|---|---|---|---|---|
| Percentage | 15% | 22% | 21% | 20% | 11% | 11% |

within each neighborhood there may be fluctuations (as also happens in reality). It is not obvious how to modify the probability contact rate between the individuals in the household depending on the size of the household. We have carried out simulations with two criteria:

a) Assuming that the probability contact rate for an individual, $j$, within household, ($W_H^j$), is the same independently of the household size.

$$W_H^j = \beta_1^{(a)} \frac{N_{\nu_{1,j}}(I_1) + N_{\nu_{1,j}}(I_2)}{N_{\nu_{1,j}} - 1} \quad (5)$$

b) Assuming that the probability contact rate for an individual within household increases proportionally to the household size

$$W_H^j = \beta_1^{(b)} \left(N_{\nu_{1,j}}(I_1) + N_{\nu_{1,j}}(I_2)\right) \quad (6)$$

In the case of households with a fixed size equal to 4, Ec. 6 is reduced to Ec. 5 if we take $\beta_1^{(b)} = \beta_1^{(a)}/3$. Then we will use: in a) $\beta_1 = 0.214$ 1/day and in b) $\beta_1 = 0.07$ 1/day, so that both cases give the same $W_H^j$ for households of size 4 as in scenario 2. For all the other parameters we take the same values as in scenario 2.

The incidences obtained for case b) are extremely similar to those of scenario 2. The only difference to mention is that in case b) the incidences corresponding to diverse samples tend to show more stochastic variations between each other. This is to be expected due to the greater spatial heterogeneity in this case.

The incidences obtained for case a) are slightly lower than those of scenario 2 and, for some samples, the rate of rise of the curve is lower and the position of the peak is shifted to the right. The agreement with the incidences of scenario 2 is recovered by slightly changing the $\beta_i$: increasing $\beta_2$ and $\beta_3$ by 10% and reducing $\beta_4$ by 14%.

In summary, given the small effect observed in these tests we have carried out, we do not expect that taking into consideration the size distribution of the households will change any of the conclusions of the study. Only a slightly greater heterogeneity in the spread of the disease could be expected.


[1] Google LLC, "Google COVID-19 community mobility reports," https://www.google.com/covid19/mobility/, accessed: 2021-02-01.

[2] Transport Ministry of Argentina, "Number of SUBE card users per day in AMBA," https://datos.transporte.gob.ar/dataset/sube-cantidad-de-tarjetas-usuarios-por-dia-en-amba, (in Spanish), accessed: 2021-10-23.

[3] Health Ministry of Argentina through the National Science Council (CONICET), part of that database is available in the following site: https://datos.gob.ar/dataset/salud-covid-19-casos-registrados-republica-argentina/archivo/salud_fd657d02-a33a-498b-a91b-2ef1a68b8d16, accessed: 2021-07-06.

[4] Ministry of Finance of Buenos Aires City Government, "COVID-19 seroprevalence survey methodology and definitive results (in Spanish)," https://www.estadisticaciudad.gob.ar/eyc/wp-content/uploads/2020/12/ir_2020_1509.pdf, acceded 2020-12-17.

[5] Ministry of Finance of Buenos Aires Province Government, "Provincial COVID-19 seroprevalence survey (in Spanish)," www.estadistica.ec.gba.gov.ar/dpe/images/INFORME_SEROPREVALENCIA.pdf, accessed: 2021-07-26.

[6] A. Jait, A. Silva, A. Bolzán, Ch. Ballejo, C. Pamparana, E. Bartel, E. García, F. Irassar, M. Pifano, M. Aguirre, M. de San Martín, M. Marro, and L. Muñoz, "Seroprevalence of antibodies against SARS-CoV-2 in popular neighbourhoods of Buenos Aires Province (in Spanish)," https://www.margen.org/pandemia/textos/barrios.pdf, accessed: 2021-02-22.

[7] Silvana Figar, Vanina Pagotto, Lorena Luna, Julieta Salto, Magdalena Wagner Manslau, Alicia S Mistchenko, Andrea Gamarnik, Ana María Gómez Saldaño, and Fernán González Bernaldo De Quirós, "Severe acute respiratory syndrome Coronavirus 2, Seroepidemiology study in Argentinian slum," MEDICINA (Buenos Aires) **81**, 135–142 (2021).

[8] Laura Muñoz, Marina Pífano, Andres Bolzán, Teresa Varela, Yamila Comes, Mariana Specogna, Leticia Ceriani, Jonatan Konfino, Nicolás Kreplak, and Enio Garcia, "Surveillance and seroprevalence: Evaluation of IgG antibodies for SARS-Cov2 by ELISA in the popular neighbourhood Villa Azul, Quilmes, Province of Buenos Aires, Argentina. Scielo Preprints 2020." https://doi.org/10.1590/SciELOPreprints.1147, in Spanish, accessed: 2021-06-17.

[9] Pedro C Hallal, Fernando P Hartwig, Bernardo L Horta, Mariângela F Silveira, Claudio J Struchiner, Luís P Vidaletti, Nelson A Neumann, Lucia C Pellanda, Odir A Dellagostin, Marcelo N Burattini, et al., "SARS-CoV-2 antibody prevalence in Brazil: results from two successive nationwide serological household surveys," The Lancet Global Health **8**, e1390–e1398 (2020).

[10] Hernan Solari, "Estimation of detectable but unde-

tected cases," https://rits.conicet.gov.ar/download/informetecnico/Apendice-4.pdf, in Spanish. Accessed: 2020-12-3.

[11] June Young Chun, Gyuseung Baek, and Yongdai Kim, "Corrigendum to Transmission onset distribution of COVID-19[Int. J. Infect. Dis. 99 (October)(2020) 403–407]," International Journal of Infectious Diseases (2021).

[12] Qun Li, Xuhua Guan, Peng Wu, Xiaoye Wang, Lei Zhou, Yeqing Tong, Ruiqi Ren, Kathy SM Leung, Eric HY Lau, Jessica Y Wong, et al., "Early transmission dynamics in Wuhan, China, of novel coronavirus–infected pneumonia," New England Journal of Medicine (2020).

[13] Wei-jie Guan, Zheng-yi Ni, Yu Hu, Wen-hua Liang, Chun-quan Ou, Jian-xing He, Lei Liu, Hong Shan, Chun-liang Lei, David SC Hui, et al., "Clinical characteristics of coronavirus disease 2019 in China," New England Journal of Medicine **382**, 1708–1720 (2020).

[14] Yongyue Wei, Liangmin Wei, Yihan Liu, Lihong Huang, Sipeng Shen, Ruyang Zhang, Jiajin Chen, Yang Zhao, Hongbing Shen, and Feng Chen, "Comprehensive estimation for the length and dispersion of COVID-19 incubation period: a systematic review and meta-analysis," Infection , 1–11 (2021).

[15] Quan-Xin Long, Xiao-Jun Tang, Qiu-Lin Shi, Qin Li, Hai-Jun Deng, Jun Yuan, Jie-Li Hu, Wei Xu, Yong Zhang, Fa-Jin Lv, et al., "Clinical and immunological assessment of asymptomatic SARS-CoV-2 infections," Nature Medicine **26**, 1200–1204 (2020).

[16] Yang Ge, Leonardo Martinez, Shengzhi Sun, Zhiping Chen, Feng Zhang, Fangyu Li, Wanwan Sun, Enfu Chen, Jinren Pan, Changwei Li, et al., "COVID-19 transmission dynamics among close contacts of index patients with COVID-19: a population-based cohort study in Zhejiang Province, China," JAMA Internal Medicine **181**, 1343–1350 (2021).

[17] Lirong Zou, Feng Ruan, Mingxing Huang, Lijun Liang, Huitao Huang, Zhongsi Hong, Jianxiang Yu, Min Kang, Yingchao Song, Jinyu Xia, et al., "SARS-CoV-2 viral load in upper respiratory specimens of infected patients," New England Journal of Medicine **382**, 1177–1179 (2020).

[18] National Center for Immunization and Respiratory Diseases (NCIRD) Division of Viral Diseases, "Ending isolation and precautions for people with COVID-19: Interim guidance," https://www.cdc.gov/coronavirus/2019-ncov/hcp/duration-isolation.html, accessed: 2021-09-14.

[19] Muge Cevik, Matthew Tate, Ollie Lloyd, Alberto Enrico Maraolo, Jenna Schafers, and Antonia Ho, "SARS-CoV-2, SARS-CoV, and MERS-CoV viral load dynamics, duration of viral shedding, and infectiousness: a systematic review and meta-analysis," The Lancet Microbe (2020).

[20] Zachary J Madewell, Yang Yang, Ira M Longini, M Elizabeth Halloran, and Natalie E Dean, "Household transmission of SARS-CoV-2: a systematic review and meta-analysis," JAMA Network Open **3**, e2031756–e2031756 (2020).

[21] PAHO Director of Communicable Diseases M.Espinal, "Virtual Press Briefing on COVID-19," https://www.facebook.com/watch/live/?ref=watch_permalink&v=656617178548546.

[22] Carlos G Grijalva, Melissa A Rolfes, Yuwei Zhu, Huong Q McLean, Kayla E Hanson, Edward A Belongia, Natasha B Halasa, Ahra Kim, Carrie Reed, Alicia M Fry, et al., "Transmission of SARS-COV-2 infections in households—Tennessee and Wisconsin, April to September 2020," Morbidity and Mortality Weekly Report **69**, 1631 (2020).

[23] Itai Dattner, Yair Goldberg, Guy Katriel, Rami Yaari, Nurit Gal, Yoav Miron, Arnona Ziv, Rivka Sheffer, Yoram Hamo, and Amit Huppert, "The role of children in the spread of COVID-19: Using household data from Bnei Brak, Israel, to estimate the relative susceptibility and infectivity of children," PLoS Computational Biology **17**, e1008559 (2021).

[24] Keeling Matt J. and Pejman Rohani, Modeling infectious diseases in humans and animals (Princeton University Press, 2008).

[25] World Health Organization, "COVID-19 natural immunity, Scientific brief, May 10th 2021," https://apps.who.int/iris/bitstream/handle/10665/341241/WHO-2019-nCoV-Sci-Brief-Natural-immunity-2021.1-eng.pdf, accessed: 2021-11-15.

[26] Argentinian National Institute of Statistics and Censuses (INDEC), "Households by number of people in the household, according to sex of head and type of household, Table H15-P, Buenos Aires Province, 2010," https://www.indec.gob.ar/ftp/censos/2010/CuadrosDefinitivos/H15-P_buenos_aires_gba.xls, in Spanish, accessed: 2021-02-19.
